# Meta-analysis fine-mapping is often miscalibrated at single-variant resolution

**DOI:** 10.1101/2022.03.16.22272457

**Authors:** Masahiro Kanai, Roy Elzur, Wei Zhou, Global Biobank Meta-analysis Initiative, Mark J Daly, Hilary K Finucane

## Abstract

Meta-analysis is pervasively used to combine multiple genome-wide association studies (GWAS) into a more powerful whole. To resolve causal variants, meta-analysis studies typically apply summary statistics-based fine-mapping methods as they are applied to single-cohort studies. However, it is unclear whether heterogeneous characteristics of each cohort (*e*.*g*., ancestry, sample size, phenotyping, genotyping, or imputation) affect fine-mapping calibration and recall. Here, we first demonstrate that meta-analysis fine-mapping is substantially miscalibrated in simulations when different genotyping arrays or imputation panels are included. To mitigate these issues, we propose a summary statistics-based QC method, SLALOM, that identifies suspicious loci for meta-analysis fine-mapping by detecting outliers in association statistics based on ancestry-matched local LD structure. Having validated SLALOM performance in simulations and the GWAS Catalog, we applied it to 14 disease endpoints from the Global Biobank Meta-analysis Initiative and found that 67% of loci showed suspicious patterns that call into question fine-mapping accuracy. These predicted suspicious loci were significantly depleted for having likely causal variants, such as nonsynonymous variants, as a lead variant (2.7x; Fisher’s exact *P* = 7.3 × 10^−4^). Compared to fine-mapping results in individual biobanks, we found limited evidence of fine-mapping improvement in the GBMI meta-analyses. Although a full solution requires complete synchronization across cohorts, our approach identifies likely spurious results in meta-analysis fine-mapping. We urge extreme caution when interpreting fine-mapping results from meta-analysis.

## Introduction

Meta-analysis is pervasively used to combine multiple genome-wide association studies (GWAS) from different cohorts^1^. Previous GWAS meta-analyses have identified thousands of loci associated with complex diseases and traits, such as type 2 diabetes^2,3^, schizophrenia^4,5^, rheumatoid arthritis^6,7^, body mass index^8^, and lipid levels^9^. These meta-analyses are typically conducted in large-scale consortia (*e*.*g*., the Psychiatric Genomics Consortium [PGC], the Global Lipids Genetics Consortium [GLGC], and the Genetic Investigation of Anthropometric Traits [GIANT] consortium) to increase sample size while harmonizing analysis plans across participating cohorts in every possible aspect (*e*.*g*., phenotype definition, quality-control [QC] criteria, statistical model, and analytical software) by sharing summary statistics as opposed to individual-level data, thereby avoiding data protection issues and variable legal frameworks governing individual genome and medical data around the world. The Global Biobank Meta-analysis Initiative (GBMI)^10^ is one such large-scale, international effort, which aims to establish a collaborative network spanning 23 biobanks from four continents (total *n* = 2.2 million) for coordinated GWAS meta-analyses, while addressing the many benefits and challenges in meta-analysis and subsequent downstream analyses.

One such challenging downstream analysis is statistical fine-mapping^11–13^. Despite the great success of past GWAS meta-analyses in locus discovery, individual causal variants in associated loci are largely unresolved. Identifying causal variants from GWAS associations (*i*.*e*., fine-mapping) is challenging due to extensive linkage disequilibrium (LD, the correlation among genetic variants), the presence of multiple causal variants, and limited sample sizes, but is rapidly becoming achievable with high confidence in individual cohorts^14–17^ owing to the recent development of large-scale biobanks^18–20^ and scalable fine-mapping methods^21–23^ that enable well-powered, accurate fine-mapping using in-sample LD from large-scale individual-level data.

After conducting GWAS meta-analysis, previous studies^2,7,9,24–30^ have applied existing summary statistics-based fine-mapping methods (*e*.*g*., approximate Bayes factor [ABF]^31,32^, CAVIAR^33^, PAINTOR^34,35^, FINEMAP^21,22^, and SuSiE^23^) just as they are applied to single-cohort studies, without considering or accounting for the unavoidable heterogeneity among cohorts (*e*.*g*. differences in sample size, phenotyping, genotyping, or imputation). Such heterogeneity could lead to false positives and miscalibration in meta-analysis fine-mapping (**Fig. 1**). For example, case-control studies enriched with more severe cases or ascertained with different phenotyping criteria may disproportionately contribute to genetic discovery, even when true causal effects for genetic liability are exactly the same between these studies and less severe or unascertained ones. Quantitative traits like biomarkers could have phenotypic heterogeneity arising from different measurement protocols and errors across studies. There might be genuine biological mechanisms too, such as gene–gene (GxG) and gene–environment (GxE) interactions and (population-specific) dominance variation (*e*.*g*., rs671 and alcohol dependence^36^), that introduce additional heterogeneity across studies^37,38^. In addition to phenotyping, differences in genotyping and imputation could dramatically undermine fine-mapping calibration and recall at single-variant resolution, because differential patterns of missingness and imputation quality across constituent cohorts of different sample sizes can disproportionately diminish association statistics of potentially causal variants. Finally, although more easily harmonized than phenotyping and genotyping data, subtle differences in QC criteria and analytical software may further exacerbate the effect of heterogeneity on fine-mapping.

**Fig. 1.**
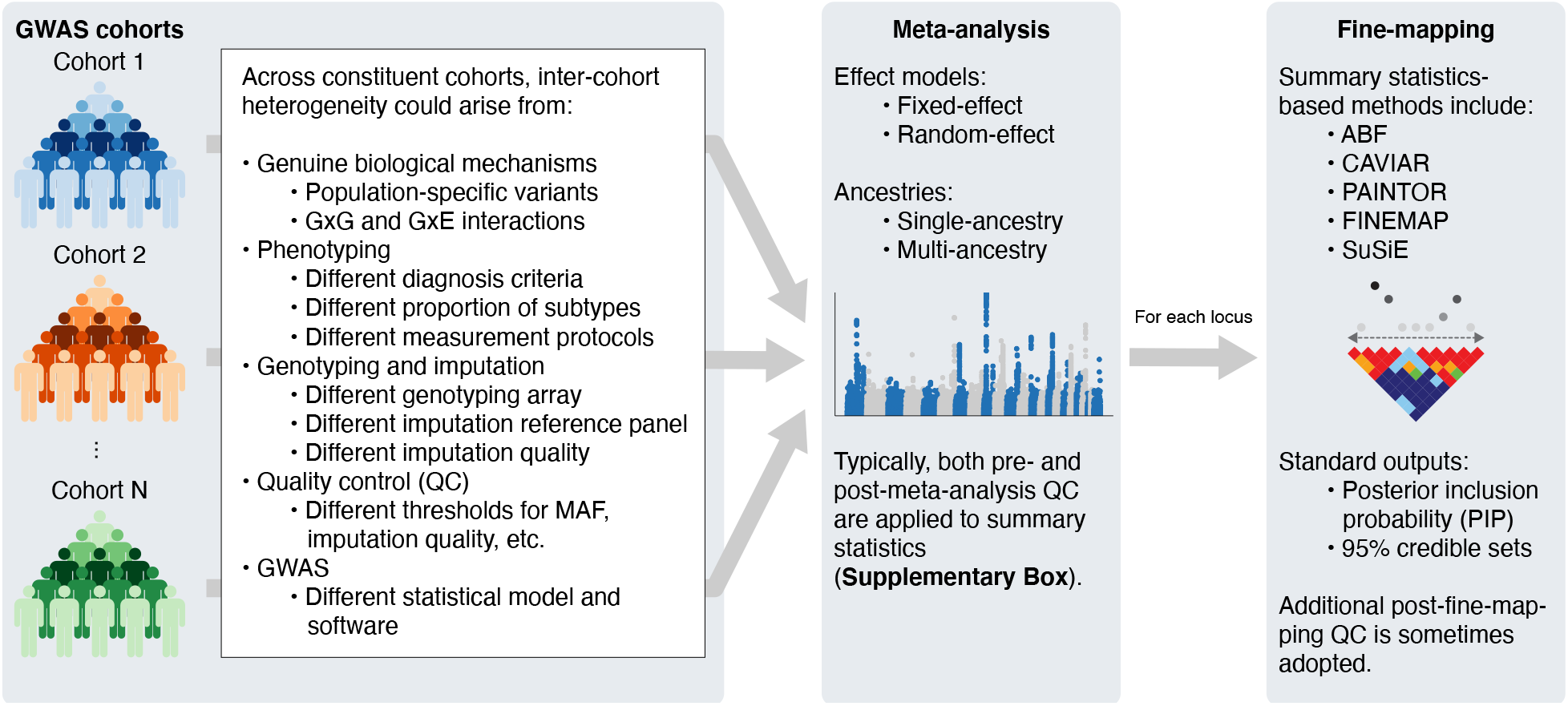
Schematic overview of meta-analysis fine-mapping.

An illustrative example of such issues can be observed in the *TYK2* locus (19p13.2) in the recent meta-analysis from the COVID-19 Host Genetics Initiative (COVID-19 HGI; **Fig. S1**)^39^. This locus is known for protective associations against autoimmune diseases^6,24^, while a complete *TYK2* loss of function results in a primary immunodeficiency^40^. Despite strong LD (*r*^2^ = 0.82) with a lead variant in the locus (rs74956615; *P* = 9.7 × 10^−12^), a known functional missense variant rs34536443 (p.Pro1104Ala) that reduces *TYK2* function^41,42^ did not achieve genome-wide significance and was assigned a very low PIP in fine-mapping (*P* = 7.5 × 10^−7^; PIP = 9.5 × 10^−4^), primarily due to its missingness in two more cohorts than rs74956615. This serves as just one example of the major difficulties with meta-analysis fine-mapping at single-variant resolution. Indeed, the COVID-19 HGI cautiously avoided an *in-silico* fine-mapping in the flagship to prevent spurious results^39^.

Only a few studies have carefully addressed these concerns in their downstream analyses. The Schizophrenia Working Group of PGC, for example, recently updated their largest meta-analysis of schizophrenia^5^ (69,369 cases and 236,642 controls), followed by a downstream fine-mapping analysis using FINEMAP^21^. Unlike many other GWAS consortia, since PGC has access to individual-level genotypes for a majority of samples, they were able to apply standardized sample and variant QC criteria and impute variants using the same reference panel, all uniformly processed using the RICOPILI pipeline^43^. This harmonized procedure was crucial for properly controlling inter-cohort heterogeneity and thus allowing more robust meta-analysis fine-mapping at single-variant resolution. Furthermore, PGC’s direct access to individual-level data enabled them to compute in-sample LD matrices for multiple causal variant fine-mapping, which prevents the significant miscalibration that results from using an external LD reference^14–16^. A 2017 fine-mapping study of inflammatory bowel disease also benefited from access to individual-level genotypes and careful pre-and post-fine-mapping QC^44^. For a typical meta-analysis consortium, however, many of these steps are infeasible as full genotype data from all cohorts is not available. For such studies, a new approach to meta-analysis fine-mapping in the presence of the many types of heterogeneity is needed. Until such a method is developed, QC of meta-analysis fine-mapping results deserves increased attention.

While existing variant-level QC procedures are effective for limiting spurious associations in GWAS (**Supplementary Box**)^45^, they do not suffice for ensuring high-quality fine-mapping results. In some cases, they even hurt fine-mapping quality, because they can i) cause or exacerbate differential patterns of missing variants across cohorts, and ii) remove true causal variants as well as suspicious variants. Thus, additional QC procedures that retain consistent variants across cohorts for consideration but limit poor-quality fine-mapping results are needed. A recently proposed method called DENTIST^46^, for example, performs summary statistics QC to improve GWAS downstream analyses, such as conditional and joint analysis (GCTA-COJO^47^), by removing variants based on estimated heterogeneity between summary statistics and reference LD. Although DENTIST was also applied prior to fine-mapping (FINEMAP^21^), simulations only demonstrated that it could improve power for detecting the correct number of causal variants in a locus, not true causal variants. This motivated us to develop a new fine-mapping QC method for better calibration and recall at single-variant resolution and to demonstrate its performance in large-scale meta-analysis.

Here, we first demonstrate the effect of inter-cohort heterogeneity in meta-analysis fine-mapping via realistic simulations with multiple heterogeneous cohorts, each with different combinations of genotyping platforms, imputation reference panels, and genetic ancestries. We propose a summary statistics-based QC method, SLALOM (<u>s</u>uspicious loci <u>a</u>nalysis <u>o</u>f <u>m</u>eta-analysis summary statistics), that identifies suspicious loci for meta-analysis fine-mapping by detecting association statistics outliers based on local LD structure, building on the DENTIST method. Applying SLALOM to 14 disease endpoints from the Global Biobank Meta-analysis Initiative^10^ as well as 467 meta-analysis summary statistics from the GWAS Catalog^48^, we demonstrate that suspicious loci for fine-mapping are widespread in meta-analysis and urge extreme caution when interpreting fine-mapping results from meta-analysis.

## Results

### Large-scale simulations demonstrate miscalibration in meta-analysis fine-mapping

Existing fine-mapping methods^21,23,31^ assume that all association statistics are derived from a single-cohort study, and thus do not model the per-variant heterogeneity in effect sizes and sample sizes that arise when meta-analyzing multiple cohorts (**Figure 1**). To evaluate how different characteristics of constituent cohorts in a meta-analysis affect fine-mapping calibration and recall, we conducted a series of large-scale GWAS meta-analysis and fine-mapping simulations (**Table S1–4**; **Methods**). Briefly, we simulated multiple GWAS cohorts of different ancestries (10 European ancestry, one African ancestry and one East Asian ancestry cohorts; *n* = 10,000 each) that were genotyped and imputed using different genotyping arrays (Illumina Omni2.5, Multi-Ethnic Global Array [MEGA], and Global Screening Array [GSA]) and imputation reference panels (the 1000 Genomes Project Phase 3 [1000GP3]^49^, the Haplotype Reference Consortium [HRC]^50^, and the TOPMed^51^). For each combination of cohort, genotyping array, and imputation panel, we conducted 300 GWAS with randomly simulated causal variants that resemble the genetic architecture of a typical complex trait, including minor allele frequency (MAF) dependent causal effect sizes^52^, total SNP heritability^53^, functional consequences of causal variants^17^, and levels of genetic correlation across cohorts (*i*.*e*., true effect size heterogeneity; *r*_g_ = 1, 0.9, and 0.5; see **Methods**). We then meta-analyzed the single-cohort GWAS results across 10 independent cohorts based on multiple *configurations* (different combinations of genotyping arrays and imputation panels for each cohort) to resemble realistic meta-analysis of multiple heterogeneous cohorts (**Table S4**). We applied ABF fine-mapping to compute a posterior inclusion probability (PIP) for each variant and to derive 95% and 99% credible sets (CS) that contain the smallest set of variants covering 95% and 99% of probability of causality. We evaluated the false discovery rate (FDR, defined as the proportion of variants with PIP > 0.9 that are non-causal) and compared against the expected proportion of non-causal variants if the meta-analysis fine-mapping method were calibrated, based on PIP. More details of our simulation pipeline are described in **Methods** and visually summarized in **Fig. S2**.

We found that FDR varied widely over the different configurations, reaching as high as 37% for the most heterogeneous configurations (**Fig. 2**). We characterized the contributing factors to the miscalibration. We first found that lower true effect size correlation *r*_g_ (*i*.*e*., larger phenotypic heterogeneity) always caused higher miscalibration and lower recall. Second, when using the same imputation panel (1000GP3), use of less dense arrays (MEGA or GSA) led to moderately inflated FDR (up to FDR = 11% vs. expected 1%), while use of multiple genotyping array did not cause further FDR inflation (**Fig. 2a**). Third, when using the same genotyping array (Omni2.5), use of imputation panels (HRC or TOPMed) that does not match our simulation reference significantly affects miscalibration (up to FDR = 17% vs. expected 1%), and using multiple imputation panels further increased miscalibration (up to FDR = 35% vs. expected 2%, **Fig. 2c**); this setup is as bad as the most heterogeneous configuration using multiple genotyping arrays and imputation panels (FDR = 37%). When TOPMed-imputed variants were lifted over from GRCh38 to GRCh37, we observed FDR increases of up to 10%, likely due to genomic build conversion failures (**Supplementary Note**)^54^. Fourth, recall was not significantly affected by heterogeneous genotyping arrays or imputation panels (**Fig. 2b,d**). Fifth, including multiple genetic ancestries did not affect calibration when using the same genotyping array and imputation panel (Omni 2.5 and 1000GP3; **Fig. 2e**) but significantly improved recall if African ancestry was included (**Fig. 2f**). This is expected, given the shorter LD length in the African population compared to other populations, which improves fine-mapping resolution^55^. Finally, in the most heterogeneous configurations where multiple genotyping arrays and imputation panels existed, we observed a FDR of up to 37% and 28% for European and multi-ancestry meta-analyses, respectively (vs. expected 2% for both), demonstrating that inter-cohort heterogeneity can substantially undermine meta-analysis fine-mapping (**Fig. 2g,h**).

**Fig. 2.**
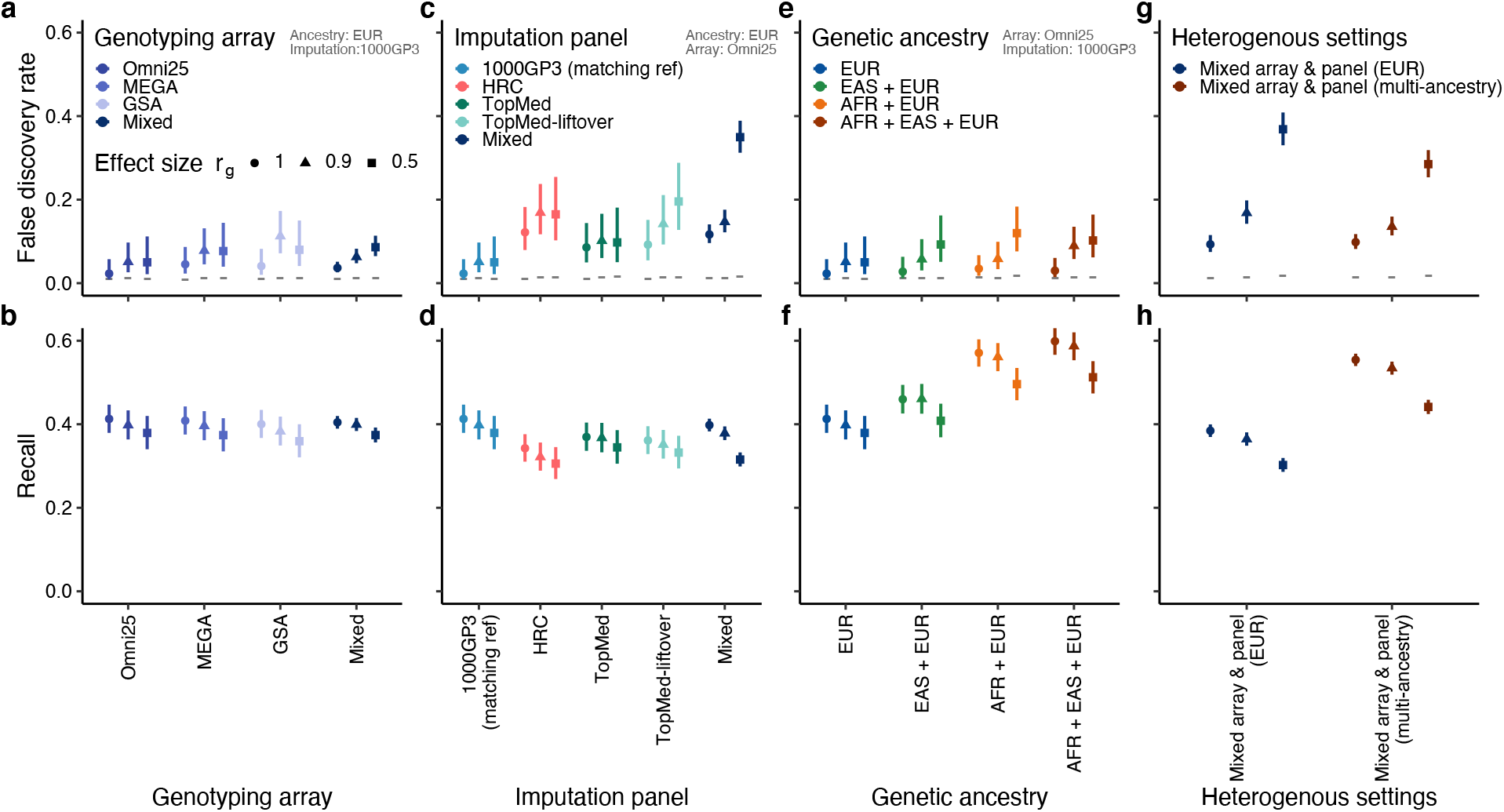
Evaluation of false discovery rate (FDR) and recall in meta-analysis fine-mapping simulations. We evaluated FDR and recall in meta-analysis fine-mapping using different genotyping arrays (**a**,**b**), imputation reference panels (**c**,**d**), genetic ancestries (**e, f**), and more heterogeneous settings by combining these (**g, h**). As shown in top-right gray labels, the EUR ancestry, the Omni2.5 genotyping array and/or the 1000GP3 reference panel were used unless otherwise stated. FDR is defined as the proportion of non-causal variants with PIP > 0.9. Horizontal gray lines represent 1 – mean PIP, *i*.*e*. expected FDR were the method calibrated. Recall is defined as the proportion of true causal variants in the top 1% PIP bin. Shapes correspond to the true effect size correlation *r*_g_ across cohorts which represent a phenotypic heterogeneity parameter (the lower *r*_g_, the higher phenotypic heterogeneity).

To further characterize observed miscalibration in meta-analysis fine-mapping, we investigated the availability of GWAS variants in each combination of ancestry, genotyping array, and imputation panel. Out of 3,285,617 variants on chromosome 3 that passed variant QC in at least one combination (per-combination MAF > 0.001 and Rsq > 0.6; **Methods**), 574,261 variants (17%) showed population-level gnomAD MAF > 0.001 in every ancestry that we simulated (African, East Asian, and European). Because we used a variety of imputation panels, we retrieved population-level MAF from gnomAD. Of these 574,261 variants, 389,219 variants (68%) were available in every combination (**Fig. S3a**). This fraction increased from 68% to 73%, 74%, and 76% as we increased gnomAD MAF thresholds to > 0.005, 0.01, and 0.05, respectively, but never reached 100% (**Fig. S4**). Notably, we observed a substantial number of variants that are unique to a certain genotyping array and an imputation panel, even when we restricted to 344,497 common variants (gnomAD MAF > 0.05) in every ancestry (**Fig. S3b**). For example, there are 34,317 variants (10%) that were imputed in the 1000GP3 and TOPMed reference but not in the HRC. Likewise, we observed 33,106 variants (10%) that were specific to the 1000GP3 reference and even 3,066 variants (1%) that were imputed in every combination except for East Asian ancestry with the GSA array and the TOPMed reference. When using different combinations of gnomAD MAF thresholds (> 0.001, 0.005, 0.01, or 0.05 in every ancestry) and Rsq thresholds (> 0.2, 0.4, 0.6, or 0.8), we observed the largest fraction of shared variants (78%) was achieved with gnomAD MAF > 0.01 and Rsq > 0.2 while the largest number of the shared variants (427,494 variants) was achieved with gnomAD MAF > 0.001 and Rsq > 0.2, leaving it unclear which thresholds would be preferable in the context of fine-mapping (**Fig. S4**).

The remaining 2,711,356 QC-passing variants in our simulations (gnomAD MAF ≤ 0.001 in at least one ancestry) further exacerbate variable coverage of the available variants (**Fig. S3c**). Of these, the largest proportion of variants (39%) were only available in African ancestry, followed by African and European (but not in East Asian) available variants (7%), European-specific variants (6%), and East Asian-specific variants (5%). Furthermore, similar to the aforementioned common variants, we found a substantial number of variants that are unique to a certain combination. Altogether, we observed that only 393,471 variants (12%) out of all the QC-passing 3,285,617 variants were available in every combination (**Fig. S3d**). These observations recapitulate that different combinations of genetic ancestry, genotyping array, imputation panels, and QC thresholds substantially affect the availability of common, well-imputed variants for association testing^56^.

Thus, the different combinations of genotyping and imputation cause each cohort in a meta-analysis to have a different set of variants, and consequently variants can have very different overall sample sizes. In our simulations with the most heterogeneous configurations, we found that 66% of the false positive loci (where a non-causal [false positive] variant was assigned PIP > 0.9) had different sample sizes for true causal and false positive variants (median maximum/minimum sample size ratio = 1.4; **Fig. S5**). Analytically, we found that at common meta-analysis sample sizes and genome-wide significant effect size regimes, when two variants have similar marginal effects, the one with the larger sample size will usually achieve a higher ABF PIP (**Supplementary Note; Fig. S6–8**). This elucidates the mechanism by which sample size imbalance can lead to miscalibration.

### Overview of the SLALOM method

To address the challenges in meta-analysis fine-mapping discussed above, we developed SLALOM (<u>s</u>uspicious <u>l</u>oci <u>a</u>nalysis <u>o</u>f <u>m</u>eta-analysis summary statistics), a method that flags suspicious loci for meta-analysis fine-mapping by detecting outliers in association statistics based on deviations from expectation, estimated with local LD structure (**Methods**). SLALOM consists of three steps, 1) defining loci and lead variants based on a 1 Mb window, 2) detecting outlier variants in each locus using meta-analysis summary statistics and an external LD reference panel, and 3) identifying suspicious loci for meta-analysis fine-mapping (**Fig. 3a,b**).

**Fig. 3.**
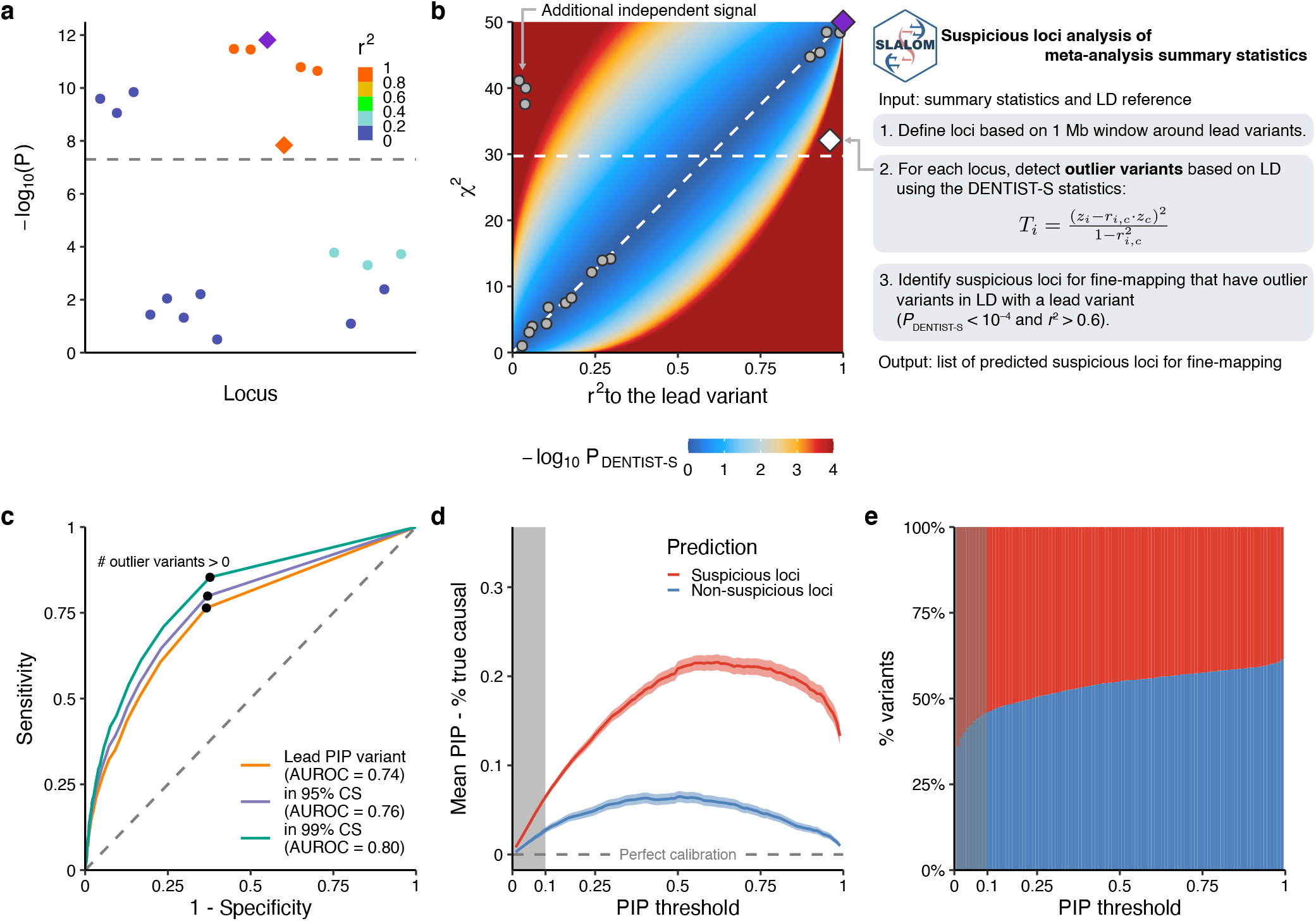
Overview of the SLALOM method. **a**,**b**. An illustrative example of the SLALOM application. **a**. In an example locus, two independent association signals are depicted: i) the most significant signal that contains a lead variant (purple diamond) and five additional variants that are in strong LD (*r*^2^ > 0.9) with the lead variant, and ii) an additional independent signal (*r*^2^ < 0.05). There is one outlier variant (orange diamond) in the first signal that deviates from the expected association based on LD. **b**. Step-by-step procedure of the SLALOM method. For outlier variant detection in a locus, a diagnosis plot of *r*^2^ values to the lead variant vs. marginal *χ*^2^ is shown to aid interpretation. Background color represents a theoretical distribution of –log_10_ *P*_DENTIST-S_ values when a lead variant has a marginal *χ*^2^ of 50, assuming no allele flipping. Points represent the variants depicted in the example locus (**a**), where the lead variant (purple diamond) and the outlier variant (white diamond) were highlighted. Diagonal line represents an expected marginal association. Horizontal dotted lines represent the genome-wide significance threshold (*P* < 5.0 × 10^−8^). **c**. The ROC curve of SLALOM prediction for identifying suspicious loci in the simulations. Positive conditions were defined as whether a true causal variant in a locus is 1) a lead PIP variant, 2) in 95% CS, and 3) in 99% CS. AUROC values were shown in the labels. Black points represent the performance of our adopted metric, *i*.*e*., whether a locus contains at least one outlier variant (*P*_DENTIST-S_ < 1.0 × 10^−4^ and *r*^2^ > 0.6). **d**. Calibration plot in the simulations under different PIP thresholds. Calibration was measured as the mean PIP – fraction of true causal variants among variants above the threshold. Shadows around the lines represent 95% confidence intervals. **e**. The fraction of variants in predicted suspicious and non-suspicious loci under different PIP thresholds. Gray shadows in the panels **d**,e reprset a PIP ≤ 0.1 regoin as we excluded loci with maximum PIP ≤ 0.1 in the actual SLALOM analysis based an this panels.

To detect outlier variants, we first assume a single causal variant per associated locus. Then the marginal z-score *z*_*i*_ for a variant *i* should be approximately equal to *r*_*i,c*_·*z*_*c*_ where *z*_*c*_ is the z-score of the causal variant *c*, and *r*_*i,c*_ is a correlation between variants *i* and *c*. For each variant in meta-analysis summary statistics, we first test this relationship using a simplified version of the DENTIST statistics^46^, DENTIST-S, based on the assumption of a single causal variant. The DENTIST-S statistics for a given variant *i* is written as

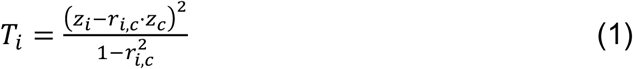

which approximately follows a *χ*^2^ distribution with 1 degree of freedom^46^. Since the true causal variant and LD structure are unknown in real data, we approximate the causal variant as the lead PIP variant in the locus (the variant with the highest PIP) and use a large-scale external LD reference from gnomAD^57^, either an ancestry-matched LD for a single-ancestry meta-analysis or a sample-size-weighted LD by ancestries for a multi-ancestry meta-analysis (**Methods**). We note that the existence of multiple independent causal variants in a locus would not affect SLALOM precision but would decrease recall (see **Discussion**).

SLALOM then evaluates whether each locus is “suspicious”—that is, has a pattern of meta-analysis statistics and LD that appear inconsistent and therefore call into question the fine-mapping accuracy. By training on loci with maximum PIP > 0.9 in the simulations, we determined that the best-performing criterion for classifying loci as true or false positives is whether a locus has a variant with *r*^2^ > 0.6 to the lead and DENTIST-S *P*-value < 1.0 × 10^−4^ (**Methods**). Using this criterion we achieved an area under the receiver operating characteristic curve (AUROC) of 0.74, 0.76, and 0.80 for identifying whether a true causal variant is a lead PIP variant, in 95% credible set (CS), and in 99% CS, respectively (**Fig. 3c**). Using different thresholds, we observed that the SLALOM performance is not very sensitive to thresholds near the threshold we chose (**Fig. S9**). We further validated the performance of SLALOM using all the loci in the simulations and observed significantly higher miscalibration in predicted suspicious loci than in non-suspicious loci (up to 16% difference in FDR at PIP > 0.9; **Fig. 3d**). We found that SLALOM-predicted “suspicious” loci tend to be from more heterogeneous configurations and the SLALOM sensitivity and specificity depends on the level of heterogeneity (**Table S5**). Given the relatively lower miscalibration and specificity at low PIP thresholds (**Fig. 3d,e**), in subsequent real data analysis we restricted the application of SLALOM to loci with maximum PIP > 0.1 (**Methods**).

### Widespread suspicious loci for fine-mapping in existing meta-analysis summary statistics

Having assessed the performance of SLALOM in simulations, we applied SLALOM to 467 meta-analysis summary statistics in the GWAS Catalog^48^ that are publicly available with a sufficient discovery sample size (*N* > 10,000; **Table S6**; **Methods**) to quantify the prevalence of suspicious loci in existing studies. These summary statistics were mostly European ancestry-only meta-analysis (63%), followed by multi-ancestry (31%), East Asian ancestry-only (3%), and African ancestry-only (2%) meta-analyses. Across 467 summary statistics from 96 publications, we identified 28,925 loci with maximum PIP > 0.1 (out of 35,864 genome-wide significant loci defined based on 1 Mb window around lead variants; **Methods**) for SLALOM analysis, of which 8,137 loci (28%) were predicted suspicious (**Table S7**).

To validate SLALOM performance in real data, we restricted our analysis to 6,065 loci that have maximum PIP > 0.1 and that contain nonsynonymous coding variants (predicted loss-of-function [pLoF] and missense) in LD with the lead variant (*r*^2^ > 0.6). Given prior evidence^16,17,44^ that such nonsynonymous variants are highly enriched for being causal, we tested the validity of our method by whether they achieve the highest PIP in the locus (*i*.*e*., successful fine-mapping) in suspicious vs. non-suspicious loci (**Methods**). While 40% (1,557 / 3,860) of non-suspicious loci successfully fine-mapped nonsynonymous variants, only 17% (384 / 2,205) of suspicious loci did, demonstrating a significant depletion (2.3x) of successfully fine-mapped nonsynonymous variants in suspicious loci (Fisher’s exact *P* = 3.6 × 10^−79^; **Fig. 4a**). We also tested whether nonsynonymous variants belonged to 95% and 99% CS and again observed significant depletion (1.4x and 1.3x, respectively; Fisher’s exact *P* < 4.6 × 10^−100^). In addition, when we used a more stringent *r*^2^ threshold (> 0.8) for selecting loci that contain nonsynonymous variants, we also confirmed significant enrichment (Fisher’s exact *P* < 6.1 × 10^−65^; **Fig. S10**). To quantify potential fine-mapping miscalibration in the GWAS Catalog, we investigated the difference between mean PIP for lead variants and fraction of lead variants that are nonsynonymous; assuming that nonsynonymous variants in these loci are truly causal, this difference equals the difference between the true and reported fraction of lead PIP variants that are causal. We observed differences between 26–51% and 10–18% under different PIP thresholds in suspicious and non-suspicious loci, respectively (**Fig. 4b**), marking 45% and 15% for high-PIP (> 0.9) variants.

**Fig. 4.**
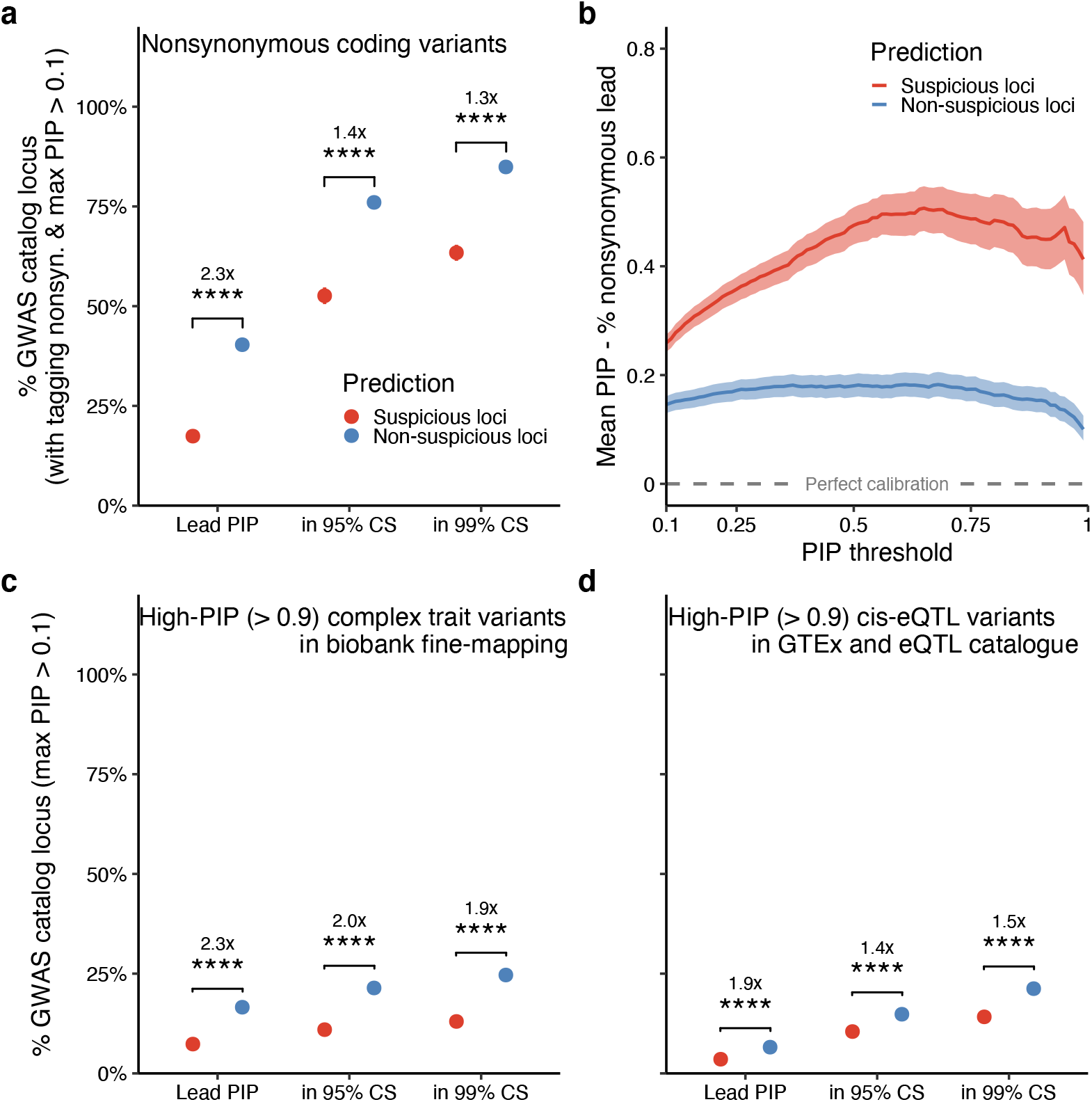
Evaluation of SLALOM performance in the GWAS Catalog summary statistics. **a**,**c**,**d**. Depletion of likely causal variants in predicted suspicious loci. We evaluated whether (**a**) nonsynonymous coding variants (pLoF and missense), (**c**) high-PIP (> 0.9) complex trait variants in biobank fine-mapping, and (**d**) high-PIP (> 0.9) *cis*-eQTL variants in GTEx v8 and eQTL Catalogue were lead PIP variants, in 95% CS, or in 99% CS in suspicious vs. non-suspicious loci. Depletion was calculated by relative risk (*i*.*e*. a ratio of proportions; **Methods**). Error bars, invisible due to their small size, correspond to 95% confidence intervals using bootstrapping. Significance represents a Fisher’s exact test *P*-value (*, *P* < 0.05; **, < 0.01; ***, < 0.001; ****, < 10^−4^). **b**. Plot of the estimated difference between true and reported proportion of causal variants in the loci tagging nonsynonymous variants (*r*^2^ > 0.6 with the lead variants) in the GWAS Catalog under different PIP thresholds. Analogous to **Fig. 3b**, assuming nonsynonymous variants in these loci are truly causal, the mean PIP for lead variants minus the fraction of lead variants that are nonsynonymous above the threshold is equal to the difference between true and reported proportion of causal variants.

We further assessed SLALOM performance in the GWAS Catalog meta-analyses by leveraging high-PIP (> 0.9) complex trait and *cis*-eQTL variants that were rigorously fine-mapped^16,17^ in large-scale biobanks (Biobank Japan [BBJ]^58^, FinnGen^20^, and UK Biobank [UKBB]^19^) and eQTL resources (GTEx^59^ v8 and eQTL Catalogue^60^). Among the 27,713 loci analyzed by SLALOM (maximum PIP > 0.1) that contain a lead variant that was included in biobank fine-mapping, 17% (3,266 / 19,692) of the non-suspicious loci successfully fine-mapped one of the high-PIP GWAS variants in biobank fine-mapping, whereas 7% (589 / 8,021) of suspicious loci did, showing a significant depletion (2.3x) of the high-PIP complex trait variants in suspicious loci (Fisher’s exact *P* = 4.6 × 10^−100^; **Fig. 4c**). Similarly, among 26,901 loci analyzed by SLALOM that contain a lead variant that was included in *cis*-eQTL fine-mapping, we found a significant depletion (1.9x) of the high-PIP *cis*-eQTL variants in suspicious loci, where 7% (1,247 / 18,976) of non-suspicious loci vs. 4% (281 / 7,925) of suspicious loci successfully fine-mapped one of the high-PIP *cis*-eQTL variants (Fisher’s exact *P* = 2.6 × 10^−24^; **Fig. 4d**). We observed the same significant depletions of the high-PIP complex trait and *cis*-eQTL variants in suspicious loci that belonged to 95% and 99% CS set (**Fig. 4c,d**).

### Suspicious loci for fine-mapping in the GBMI summary statistics

Next, we applied SLALOM to meta-analysis summary statistics of 14 disease endpoints from the GBMI^10^. These summary statistics were generated from a meta-analysis of up to 1.8 million individuals in total across 18 biobanks for discovery, representing six different genetic ancestry groups of approximately 33,000 African, 18,000 Admixed American, 31,000 Central and South Asian, 341,000 East Asian, 1.4 million European, and 1,600 Middle Eastern individuals (**Table S8**). Among 489 genome-wide significant loci across the 14 traits (excluding the major histocompatibility complex [MHC] region, **Methods**), we found that 82 loci (17%) showed maximum PIP < 0.1, thus not being further considered by SLALOM. Of the remaining 407 loci with maximum PIP > 0.1, SLALOM identified that 272 loci (67%) were suspicious loci for fine-mapping (**Fig. 5a**; **Table S9**). The fraction of suspicious loci and their maximum PIP varied by trait, reflecting different levels of statistical power (*e*.*g*., sample sizes, heritability, and local LD structure) as well as inter-cohort heterogeneity (**Fig. 5b–o**).

**Fig. 5.**
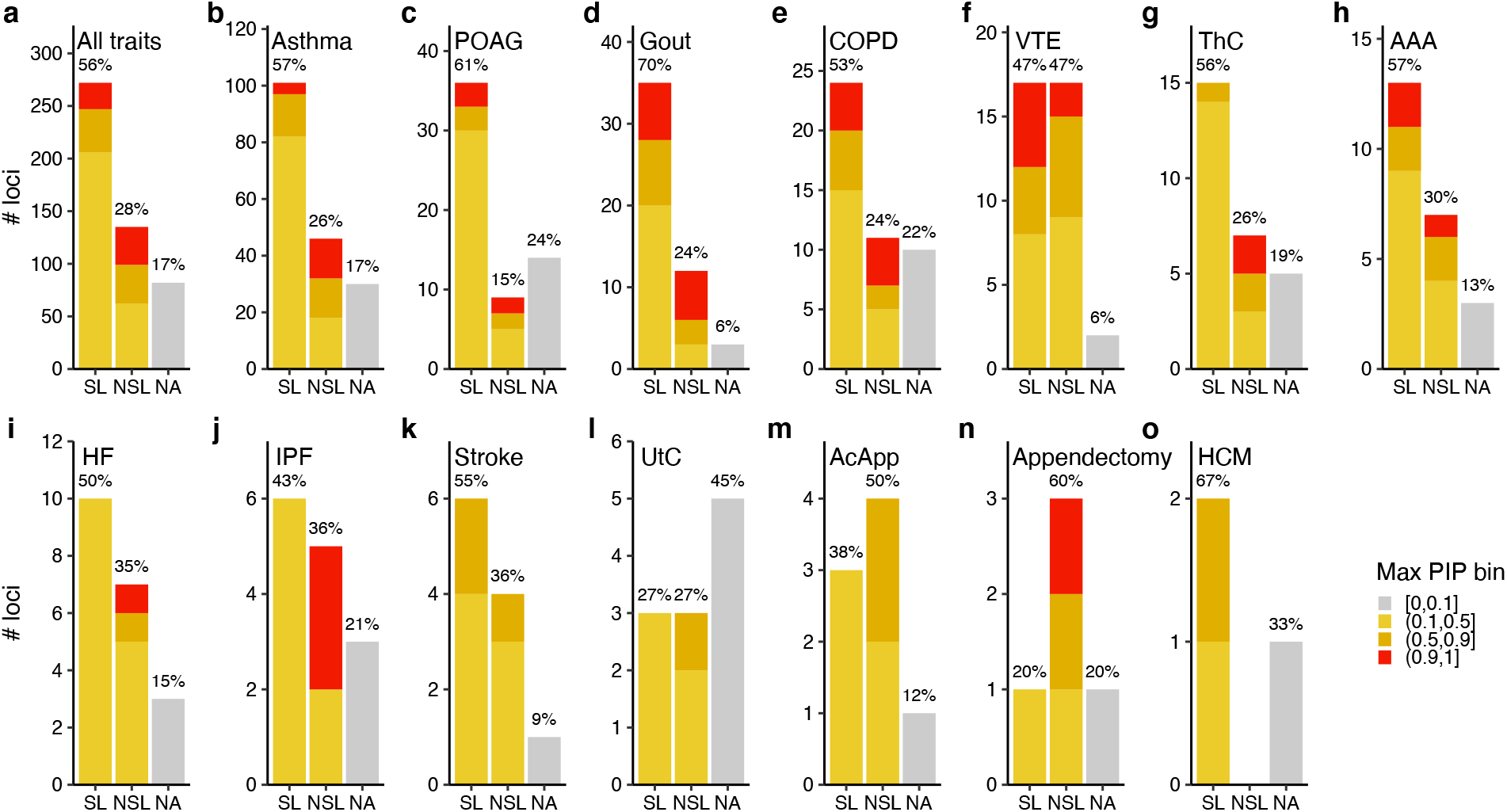
SLALOM prediction results in the GBMI summary statistics. For (**a**) all 14 traits and (**b–o**) individual traits, a number of predicted suspicious (SL), non-suspicious (NSL), and non-applicable (NA; maximum PIP < 0.1) loci were summarized. Individual traits are ordered by the total number of loci. Color represents the maximum PIP in a locus. Label represents the fraction of loci in each prediction category. AAA, abdominal aortic aneurysm. AcApp, acute appendicitis. COPD, chronic obstructive pulmonary disease. HCM, hypertrophic cardiomyopathy. HF, heart failure. IPF, idiopathic pulmonary fibrosis. POAG, primary open angle glaucoma. ThC, thyroid cancer. UtC, uterine cancer. VTE, venous thromboembolism.

While the fraction of suspicious loci (67%) in the GBMI meta-analyses is higher than in the GWAS Catalog (28%), there might be multiple reasons for this discrepancy, including association significance, sample size, ancestral diversity, and study-specific QC criteria. For example, the GBMI summary statistics were generated from multi-ancestry, large-scale meta-analyses of median sample size of 1.4 million individuals across six ancestries, while 63% of the 467 summary statistics from the GWAS Catalog were only in European-ancestry studies and 83% had less than 0.5 million discovery samples. Nonetheless, predicted suspicious loci for fine-mapping were prevalent in both the GWAS Catalog and the GBMI.

Using nonsynonymous (pLoF and missense) and high-PIP (> 0.9) complex trait and *cis*-eQTL variants, we recapitulated a significant depletion of these likely causal variants in predicted suspicious loci (2.7x, 5.2x, and 5.1x for nonsynonymous, high-PIP complex trait, and high-PIP *cis*-eQTL variants being a lead PIP variant, respectively; Fisher’s exact *P* < 7.3 × 10^−4^), confirming our observation in the GWAS Catalog analysis (**Fig. 6a–c**).

**Fig. 6.**
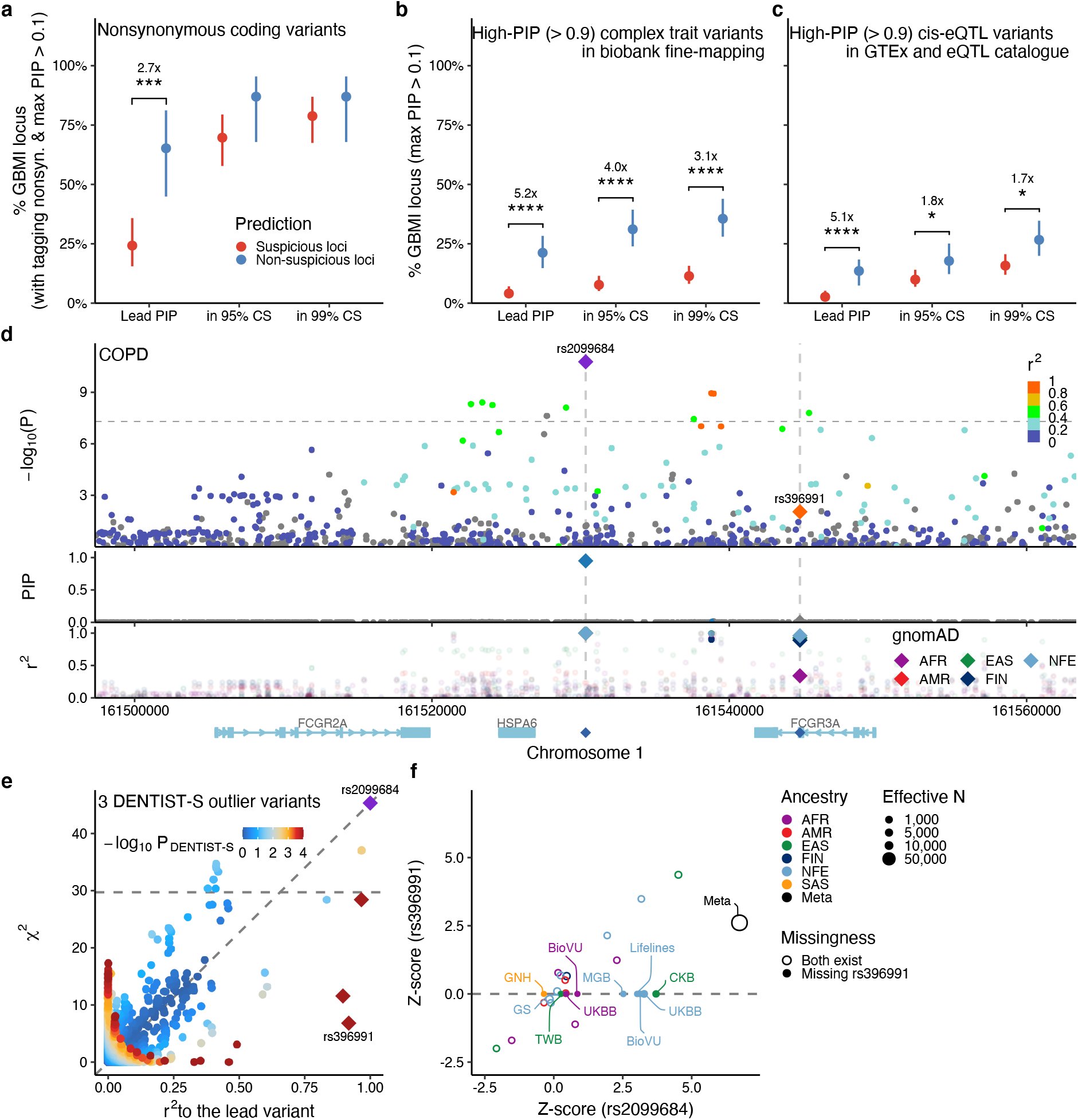
Evaluation of SLALOM performance in the GBMI summary statistics. **a–c**. Similar to **Fig. 4**, we evaluated whether (**a**) nonsynonymous coding variants (pLoF and missense), (**b**) high-PIP (> 0.9) complex trait variants in biobank fine-mapping, and (**c**) high-PIP (> 0.9) *cis*-eQTL variants in GTEx v8 and eQTL Catalogue were lead PIP variants, in 95% CS, or in 99% CS in suspicious vs. non-suspicious loci. Depletion was calculated by relative risk (*i*.*e*. a ratio of proportions; **Methods**). Error bars correspond to 95% confidence intervals using bootstrapping. Significance represents a Fisher’s exact test *P*-value (*, *P* < 0.05; **, < 0.01; ***, < 0.001; ****, < 10^−4^). **d**. Locuszoom plot of the 1q23.3 locus for COPD. The top panel shows a Manhattan plot, where the lead variant rs2099684 (purple diamond) and a missense variant rs396991 (orange diamond) are highlighted. Color represents *r*^2^ values to the lead variant. Horizontal line represents a genome-wide significance threshold (*P* = 5.0 × 10^−8^). The middle panel shows PIP from ABF fine-mapping. Color represents whether variants belong to a 95% CS. The bottom panel shows *r*^2^ values with the lead variant in gnomAD populations. **e**. A diagnosis plot showing *r*^2^ values to the lead variant vs. marginal *χ*^2^. Color represents –log_10_ *P*_DENTIST-S_ values. Outlier variants with *P*_DENTIST-S_ < 10^−4^ are depicted in red with a diamond shape. Diagonal line represents an expected marginal association. Horizontal line represents a genome-wide significance threshold. f. Z-scores of the lead variant (rs2099684) vs. the missense variant (rs396991) in the constituent cohorts of the meta-analysis. Open and closed circles represent whether both variants exist in a cohort or rs396991 is missing. Circle size corresponds to an effective sample size. Color represents genetic ancestry.

In 15/23 non-suspicious loci harboring a nonsynonymous variant, the nonsynonymous variant had the highest PIP. These included known missense variants such as rs116483731 (p.Arg20Gln) in *SPDL1* for idiopathic pulmonary fibrosis (IPF)^61,62^ and rs28929474 (p.Glu366Lys) in *SERPINA1* for chronic obstructive pulmonary disease (COPD)^63,64^. In addition, we observed successful fine-mapping in 2 novel loci for asthma, i) rs41286560 (p.Pro558Thr) in *RTL1*, a missense variant known for decreasing height^65,66^ and ii) rs34187696 (p.Gly337Val) in *ZSCAN5A*, a known missense variant for increasing monocyte count^30^.

To characterize fine-mapping failures in suspicious loci, we examined suspicious loci in which a nonsynonymous variant did not achieve the highest PIP. For example, the *FCGR2A*/*FCGR3A* (1q23.3) locus for COPD contained a genome-wide significant lead intergenic variant rs2099684 (*P* = 1.7 × 10^−11^) which is in LD (*r*^2^ = 0.92) with a missense variant rs396991 (p.Phe176Val) of *FCGR3A* (**Fig. 6d**). This locus was not previously reported for COPD, but is known for associations with autoimmune diseases (*e*.*g*., inflammatory bowel disease^44^, rheumatoid arthritis^7^, and systemic lupus erythematosus^67^) and encodes the low-affinity human FC-gamma receptors that bind to the Fc region of IgG and activate immune responses^68^. Notably, this locus contains copy number variations that contribute to the disease associations in addition to single-nucleotide variants, which makes genotyping challenging^68,69^. Despite strong LD with the lead variant, rs396991 did not achieve genome-wide significance (*P* = 9.1 × 10^−3^), showing a significant deviation from the expected association (*P*_DENTIST-S_ = 5.3 × 10^−41^; **Fig. 6e**). This is primarily due to missingness of rs396991 in 8 biobanks out of 17 (*N*_eff_ = 76,790 and 36,781 for rs2099684 and rs396991, respectively; **Fig. 6f**), which is caused by its absence from major imputation reference panels (*e*.*g*., 1000GP^49^, HRC^50^, and UK10K^70^) despite having a high MAF in every population (MAF = 0.24–0.34 in African, admixed American, East Asian, European, and South Asian populations of gnomAD^57^).

Sample size imbalance across variants was pervasive in the GBMI meta-analyses^71^, and was especially enriched in predicted suspicious loci—84% of suspicious loci vs. 24% of non-suspicious loci showed a maximum/minimum effective sample size ratio > 2 among variants in LD (*r*^2^ > 0.6) with lead variants (a median ratio = 4.2 and 1.2 in suspicious and non-suspicious loci, respectively; **Fig. S11**). These observations are consistent with our simulations, recapitulating that sample size imbalance results in miscalibration for meta-analysis fine-mapping. Notably, we observed a similar issue in other GBMI downstream analyses (*e*.*g*., polygenic risk score [PRS]^71^ and drug discovery^72^), where predictive performance improved significantly after filtering out variants with maximum *N*_eff_ < 50%. Although fine-mapping methods cannot simply take this approach because it inevitably reduces calibration and recall by removing true causal variants, other meta-analysis downstream analyses that primarily rely on polygenic signals rather than individual variants should consider this filtering as an extra QC step.

### Comparison of fine-mapping results between the GBMI meta-analyses and individual biobanks

Motivated by successful validation of SLALOM performance, we investigated whether fine-mapping confidence and resolution were improved in the GBMI meta-analyses over individual biobanks. To this end, we used our fine-mapping results^16,17^ of nine disease endpoints (asthma^64^, COPD^64^, gout, heart failure^73^, IPF^62^, primary open angle glaucoma^74^, thyroid cancer, stroke^75^, and venous thromboembolism^76^) in BBJ^58^, FinnGen^20^, and UKBB^19^ Europeans that also contributed to the GBMI meta-analyses for the same traits.

To perform an unbiased comparison of PIP between the GBMI meta-analysis and individual biobanks, we investigated functional enrichment of fine-mapped variants based on top PIP rankings in the GBMI and individual biobanks (top 0.5%, 0.1%, and 0.05% PIP variants in the GBMI vs. maximum PIP across BBJ, FinnGen, and UKBB; **Methods**). Previous studies have shown that high-PIP (> 0.9) complex trait variants are significantly enriched for well-known functional categories, such as coding (pLoF, missense, and synonymous), 5’/3’ UTR, promoter, and *cis*-regulatory element (CRE) regions (DNase I hypersensitive sites [DHS] and H3K27ac)^16,17^. Using these functional categories, we found no significant enrichment of variants in the top PIP rankings in the GBMI over individual biobanks (Fisher’s exact *P* > 0.05; **Fig. 7a**) except for variants in the promoter region (1.8x; Fisher’s exact *P* = 4.9 × 10^−4^ for the top 0.1% PIP variants). We observed similar trends regardless of whether variants were in suspicious or non-suspicious loci (**Fig. 7b,c**). To examine patterns of increased and decreased PIP for individual variants, we also calculated PIP difference between the GBMI and individual biobanks, defined as ΔPIP = PIP (GBMI) – maximum PIP across BBJ, FinnGen, and UKBB (**Fig. S12**,**13)**. We investigated functional enrichment based on ΔPIP bins and observed inconsistent enrichment results using different ΔPIP thresholds (**Fig. S14**). Finally, to test whether fine-mapping resolution was improved in the GBMI over individual biobanks, we compared the size of 95% CS after restricting them to cases where a GBMI CS overlapped with an individual biobank CS from BBJ, FinnGen, or UKBB (**Methods**). We observed the median 95% CS size of 2 and 2 in non-suspicious loci for the GBMI and individual biobanks, respectively, and 5 and 14 in suspicious loci, respectively (**Fig. S15**). The smaller credible set size in suspicious loci in GBMI could be due to improved resolution or to increased miscalibration. These results provide limited evidence of overall fine-mapping improvement in the GBMI meta-analyses over what is achievable by taking the best result from individual biobanks.

**Fig. 7.**
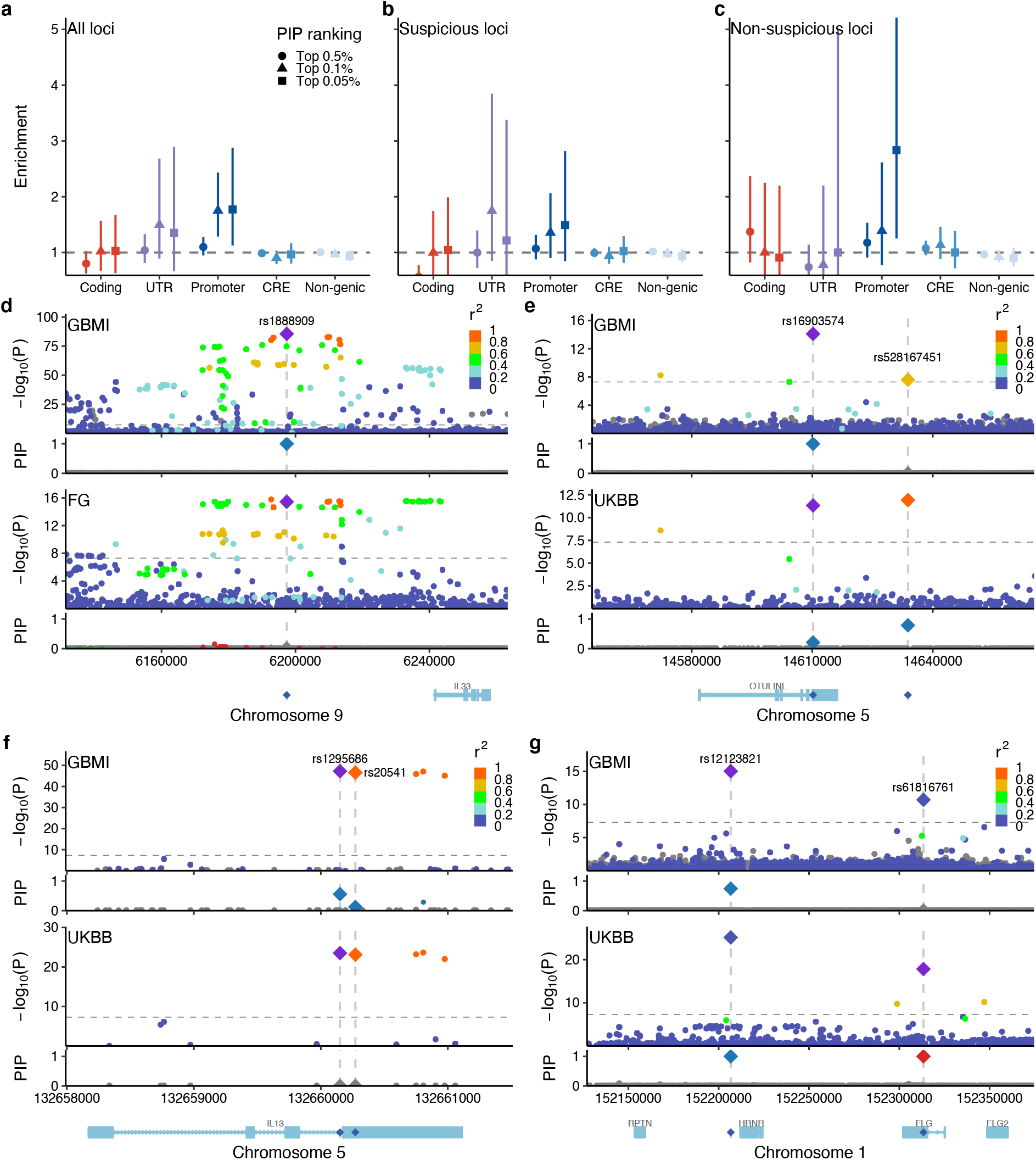
Fine-mapping improvement and retrogression in the GBMI meta-analyses over individual biobanks. a– c. Functional enrichment of variants in each functional category based on top PIP rankings in the GBMI and individual biobanks (maximum PIP of BBJ, FinnGen, and UKBB). Shape corresponds to top PIP ranking (top 0.5%, 0.1%, and 0.05%). Enrichment was calculated by a relative risk (*i.e*. a ratio of proportions; **Methods**). Error bars correspond to 95% confidence intervals using bootstrapping. **d–g**. Locuszoom plots for the same non-suspicious locus of asthma in the GBMI meta-analysis and an individual biobank (BBJ, FinnGen, or UKBB Europeans) that showed the highest PIP in our biobank fine-mapping. Colors in the Manhattan panels represent r2 values to the lead variant. In the PIP panels, only fine-mapped variants in the 95% CS are colored, where the same colors are applied between the GBMI meta-analysis and an individual biobank based on merged CS as previously described. Horizontal line represents a genome-wide significance threshold (*P* = 5.0 × 10^−8^). **d**. rs1888909 for asthma in the GBMI and FinnGen. **e**. rs16903574 for asthma in the GBMI and UKBB Europeans. Nearby rs528167451 was also highlighted, which was in strong LD (*r*^2^ = 0.86) and in the same 95% CS in UKBB Europeans, but not in the GBMI (*r*^2^ = 0.67). **f**. rs1295686 for asthma in the GBMI and UKBB Europeans. A nearby missense, rs20541, showed lower PIP than rs1295686 despite having strong LD (*r*^2^ = 0.96). **g**. rs12123821 for asthma in the GBMI and UKBB Europeans. Nearby stop-gained rs61816761 was independent of rs12123821 (*r*^2^ = 0.0) and not fine-mapped in the GBMI due to a single causal variant assumption in the ABF fine-mapping.

Individual examples, however, provide insights into the types of fine-mapping differences that can occur. To characterize the observed differences in fine-mapping confidence and resolution, we further examined non-suspicious loci with ΔPIP > 0.5 in asthma. In some cases, the increased power and/or ancestral diversity of GBMI led to improved fine-mapping: for example, an intergenic variant rs1888909 (∼18 kb upstream of *IL33*) showed ΔPIP = 0.99 (PIP = 1.0 and 0.008 in GBMI and FinnGen, respectively; **Fig. 7d**), which was primarily owing to increased association significance in a meta-analysis (*P* = 3.0 × 10^−86^, 7.4 × 10^−2^, 3.6 × 10^−16^, and 1.9 × 10^−53^ in GBMI, BBJ, FinnGen, and UKBB Europeans, respectively) as well as a shorter LD length in the African population than in the European population (LD length = 4 kb vs. 41 kb for variants with *r*^2^ > 0.6 with rs1888909 in the African and European populations, respectively; *N*_eff_ = 4,270 for Africans in the GBMI asthma meta-analysis; **Fig. S16**). This variant was also fine-mapped for eosinophil count in UKBB Europeans (PIP = 1.0; *P* = 1.3 × 10^−314^)^16^ and was previously reported to regulate *IL33* gene expression in human airway epithelial cells via allele-specific transcription factor binding of OCT-1 (POU2F1)^77^. Likewise, we observed a missense variant rs16903574 (p.Phe319Leu) in *OTULINL* showed ΔPIP = 0.79 (PIP = 1.0 and 0.21 in GBMI and UKBB

Europeans, respectively; **Fig. 7e**) owing to improved association significance (*P* = 7.7 × 10^−15^ and 4.7 × 10^−12^ in GBMI and UKBB Europeans, respectively).

However, we also observed very high ΔPIP for variants that are not likely causal. For example, we observed that an intronic variant rs1295686 in *IL13* showed ΔPIP = 0.56 (PIP = 0.56 and 0.0002 in GBMI and UKBB Europeans, respectively; **Fig. 7f**), despite having strong LD with a nearby missense variant rs20541 (p.Gln144Arg; *r*^2^ = 0.96 with rs1295686) which only showed ΔPIP = 0.13 (PIP = 0.13 and 0.0001 in GBMI and UKBB Europeans, respectively). The missense variant rs20541 showed PIP = 0.23 and 0.15 for a related allergic disease, atopic dermatitis, in BBJ and FinnGen, respectively^17^, and was previously shown to induce STAT6 phosphorylation and up-regulate CD23 expression in monocytes, promoting IgE synthesis^78^. Although the GBMI meta-analysis contributed to prioritizing these two variants (sum of PIP = 0.69 vs. 0.0003 in GBMI and UKBB Europeans, respectively), the observed ΔPIP was higher for rs1295686 than for rs20541.

While increasing sample size in meta-analysis improves association significance, we also found negative ΔPIP due to losing the ability to model multiple causal variants. A stop-gained variant rs61816761 (p.Arg501Ter) in *FLG* showed ΔPIP = –1.0 (PIP = 6.4 × 10^−5^ and 1.0 in GBMI and UKBB Europeans, respectively; **Fig. 7g**), which was primarily owing to a nearby lead variant rs12123821 (∼17 kb downstream of *HRNR*; *r*^2^ = 0.0 with rs61816761). This lead variant rs12123821 showed greater significance than rs61816761 in GBMI (*P* = 9.3 × 10^−16^ and 2.0 × 10^− 11^ for rs12123821 and rs61816761, respectively) as well as in UKBB Europeans (*P* = 7.1 × 10^−26^ and 1.5 × 10^−18^). While our biobank fine-mapping^16,17^ assigned PIP = 1.0 for both variants based on multiple causal variant fine-mapping (*i*.*e*., FINEMAP^21^ and SuSiE^23^), our ABF fine-mapping in the GBMI meta-analysis was only able to assign PIP = 0.74 for the lead variant rs12123821 due to a single causal variant assumption. This recapitulates the importance of multiple causal variant fine-mapping in complex trait fine-mapping^16,17^—however, we note that multiple causal variant fine-mapping with an external LD reference is extremely error-prone as previously reported^14–16^.

## Discussion

In this study, we first demonstrated in simulations that meta-analysis fine-mapping is substantially miscalibrated when constituent cohorts are heterogeneous in phenotyping, genotyping, and imputation. To mitigate this issue, we developed SLALOM, a summary statistics-based QC method for identifying suspicious loci in meta-analysis fine-mapping. Applying SLALOM to 14 disease endpoints from the GBMI meta-analyses^10^ as well as 467 summary statistics from the GWAS Catalog^48^, we observed widespread suspicious loci in meta-analysis summary statistics, suggesting that meta-analysis fine-mapping is often miscalibrated in real data too. Indeed, we demonstrated that the predicted suspicious loci were significantly depleted for having likely causal variants as a lead PIP variant, such as nonsynonymous variants, high-PIP (> 0.9) GWAS and cis-eQTL fine-mapped variants from our previous fine-mapping studies^16,17^. Our method provides better calibration in non-suspicious loci for meta-analysis fine-mapping, generating a more reliable set of variants for further functional characterization.

We have found limited evidence of improved fine-mapping in the GBMI meta-analyses over individual biobanks. A few empirical examples in this study as well as other previous studies^7,9,26,27,30^ suggested that multi-ancestry, large-scale meta-analysis could have potential to improve fine-mapping confidence and resolution owing to increased statistical power in associations and differential LD pattern across ancestries. However, we have highlighted that the observed improvement in PIP could be due to sample size imbalance in a locus, miscalibration, and technical confoundings too, which further emphasizes the importance of careful investigation of fine-mapped variants identified through meta-analysis fine-mapping. Given practical challenges in data harmonization across different cohorts, a large-scale biobank with multiple ancestries (*e*.*g*., UK Biobank^19^ and All of Us^79^) would likely benefit the most from meta-analysis fine-mapping across ancestries.

As high-confidence fine-mapping results in large-scale biobanks and molecular QTLs continue to become available^16,17,60^, we propose alternative approaches for prioritizing candidate causal variants in a meta-analysis. First, these high-confidence fine-mapped variants have been a valuable resource to conduct a “PheWAS”^16^ to match with associated variants in a meta-analysis, which provides a narrower list of candidate variants assuming they would equally be functional and causal in related complex traits or tissues/cell-types. Second, a traditional approach based on tagging variants (*e*.*g*., *r*^2^ > 0.6 with lead variants, or PICS^80^ fine-mapping approach that only relies on a lead variant and LD) can be still highly effective, especially for known functional variants such as nonsynonymous coding variants. As we highlighted in this and previous^39^ studies, potentially causal variants in strong LD with lead variants might not achieve genome-wide significance because of missingness and heterogeneity.

While using an external LD reference for fine-mapping has been shown to be extremely error-prone^14–16^, we find here that it can be useful for flagging suspicious loci, even when it does not perfectly represent the in-sample LD structure of the meta-analyzed individuals. However, our use of external LD reference comes with several limitations. For example, due to the finite sample size of external LD reference, rare or low-frequency variants have larger uncertainties around *r*^2^ than common variants. Moreover, our *r*^2^ values in a multi-ancestry meta-analysis are currently approximated based on a sample-size-weighted average of *r*^2^ across ancestries as previously suggested^81^, but this can be different from actual *r*^2^. These uncertainties around *r*^2^ affect SLALOM prediction performance and should be modeled appropriately for further method development. On the other hand, we find it challenging to use a LD reference when true causal variants are located within a complex region (*e*.*g*., major histocompatibility complex [MHC]), or are entirely missing from standard LD or imputation reference panels, especially for structural variants. These limitations are not specific to meta-analysis fine-mapping, and separate fine-mapping methods based on bespoke imputation references have been developed (*e*.*g*., HLA^82^, KIR^83^, and variable numbers of tandem repeats [VNTR]^84^).

In addition, there are several methodological limitations of SLALOM. First, our simulations only include one causal variant per locus. Although additional independent causal variants would not affect SLALOM precision (but decrease recall), multiple *correlated* causal variants in a locus would violate SLALOM assumptions and could lead to some DENTIST-S outliers that are not due to heterogeneity or missingness but rather simply a product of tagging multiple causal variants in LD. In fact, our previous studies have illustrated infrequent but non-zero presence of such correlated causal variants in complex traits^16,17^. Second, SLALOM prediction is not perfect. Although fine-mapping calibration is certainly better in non-suspicious vs. suspicious loci, SLALOM has low precision, and we still observe some miscalibration in non-suspicious loci. Optimal thresholds for SLALOM prediction might be different for other datasets. Third, SLALOM does not model effect size heterogeneity. Although SLALOM is able to detect suspicious loci due to effect size heterogeneity as the method is agnostic to the source of heterogeneity, methods which model effect size heterogeneity, such as MR-MEGA^85^, could improve SLALOM performance. Finally, SLALOM is a per-locus QC method and does not calibrate per-variant PIPs. Further methodological development that properly models heterogeneity, missingness, sample size imbalance, multiple causal variants, and LD uncertainty across multiple cohorts and ancestries is needed to refine per-variant calibration and recall in meta-analysis fine-mapping.

We have found evidence in our simulations and real data of severe miscalibration of fine-mapping results from GWAS meta-analysis; for example, we estimate that the difference between true and reported proportion of causal variants is 20% and 45% for high-PIP (> 0.9) variants in suspicious loci from the simulations and the GWAS Catalog, respectively. Our SLALOM method helps to exclude spurious results from meta-analysis fine-mapping; however, even fine-mapping results in SLALOM-predicted “non-suspicious” loci remain somewhat miscalibrated, showing estimated differences between true and reported proportion of causal variants of 4% and 15% for high-PIP variants in the simulations and the GWAS Catalog, respectively. We thus urge extreme caution when interpreting PIPs computed from meta-analyses until improved methods are available. We recommend that researchers looking to identify likely causal variants employ complete synchronization of study design, case/control ascertainment, genomic profiling, and analytical pipeline, or rely more heavily on functional annotations, biobank fine-mapping, or molecular QTLs.

## Data Availability

The GBMI summary statistics for the 14 endpoints are available at https://www.globalbiobankmeta.org/resources and are browserble at the GBMI PheWeb website (http://results.globalbiobankmeta.org/).

http://results.globalbiobankmeta.org/

## Acknowledgements

We acknowledge all the participants and researchers of the 23 biobanks that have contributed to the GBMI. Biobank-specific acknowledgements are included in the **Supplementary Note**. We thank H. Huang, A.R. Martin, B.M. Neale, Y. Okada, K. Tsuo, J.C. Ulirsch, Y. Wang, and all the members of Finucane and Daly labs for their helpful feedback. M.K. was supported by a Nakajima Foundation Fellowship and the Masason Foundation. H.K.F. was funded by NIH grant DP5 OD024582.

## Author contributions

M.K., M.J.D, and H.K.F. designed the study. M.K., R.E. and W.Z. performed analyses. H.K.F supervised this work. H.K.F. and M.K. obtained funding. M.K., R.E., M.J.D., and H.K.F. wrote the manuscript with input from all authors.

## STAR Methods Resource availability

### Lead contact

Further information and requests for resources and data should be directed to and will be fulfilled by the lead contact, Masahiro Kanai.

### Materials availability

This study did not generate new unique reagents.

### Data and code availability

The GBMI summary statistics for the 14 endpoints are publicly available and are browserble at the GBMI PheWeb website (http://results.globalbiobankmeta.org/). Example outputs from the meta-analysis fine-mapping simulation pipeline have been deposited at Harvard Dataverse. All original code has been deposited at Zenodo and is publicly available as of the date of publication. DOIs and links are listed in the key resources table. Any additional information required to reanalyze the data reported in this paper is available from the lead contact upon request.

## Method details

### Meta-analysis fine-mapping simulation

To benchmark fine-mapping performance in meta-analysis, we simulated a large-scale, realistic GWAS meta-analysis and performed fine-mapping under different scenarios. An overview of our simulation pipeline is summarized in **Fig. S2**.

#### Simulated true genotype

Using HAPGEN2^87^ with the 1000 Genomes Project Phase 3 reference^49^, we simulated “true” genotypes of chromosome 3 for multiple independent cohorts from African, East Asian, and European ancestries. For each independent cohort from a given ancestry, we simulated 10,000 individuals each using the default parameters, with an ancestry-specific effective population size set to 17,469, 14,269, and 11,418 for Africans, East Asians, and Europeans, respectively, as recommended^87^. To mimic sample size imbalance of different ancestries in the current meta-analyses, we simulated 10 independent European cohorts, 1 African cohort, and 1 East Asian cohort.

To restrict our analysis to unrelated samples, we computed sample relatedness based on KING kinship coefficients^88^ using PLINK 2.0 (ref. ^89^) and removed monozygotic twins, duplicated individuals, or first-degree relatives with the coefficient threshold of 0.177. The detailed sample sizes of unrelated individuals for each cohort is summarized in **Table S1**.

#### Genotyping and imputation

To simulate realistic genotyping and imputation procedures, we first virtually genotyped each cohort by restricting variants to those that are available on different genotyping arrays. We selected three major genotyping arrays from Illumina, Inc. (Omni2.5, Multi-Ethnic Global Array [MEGA], and Global Screening Array [GSA]) that have different densities of genotyping probes (**Table S2**). For each cohort, we created three virtually genotyped datasets by retaining variants that are genotyped on each array. For the sake of simplicity, we assumed no genotyping errors occurred between true genotypes and virtually genotyped data—however, in practice, genotyping error is one of the major sources of unexpected confounding (*e*.*g*., see recent discussions here^90,91^) and should be treated carefully.

For each pair of cohort and genotyping array, we then imputed missing variants using different imputation reference panels. We used the Michigan Imputation Server (https://imputationserver.sph.umich.edu/)^92^ and the TOPMed Imputation Server (https://imputation.biodatacatalyst.nhlbi.nih.gov/)^51^ with the default parameters, using three publicly available reference panels: the 1000 Genomes Project Phase 3 (version 5; *n* = 2,504; 1000GP3)^49^, the Haplotype Reference Consortium (version r1.1; *n* = 32,470; HRC)^50^, and the TOPMed (version R2; n = 97,256)^51^. Briefly, for each input, the imputation server created chunks of 20 Mb, applied the standard QC, pre-phased each chunk with Eagle2 (ref. ^93^), and imputed non-genotyped variants using a specified reference panel with Minimac4 (https://genome.sph.umich.edu/wiki/Minimac4). The detailed documentation of the imputation pipeline is available on the Michigan and TOPMed websites and has been described elsewhere^92^.

We applied post-imputation QC by only keeping variants with MAF > 0.001 and imputation Rsq > 0.6. Because the TOPMed panel is based on GRCh38 while the 1000GP3 and the HRC panels are on GRCh37, we lifted over TOPMed variants from GRCh38 to GRCh37 to meta-analyze with other cohorts. We excluded any variants which were lifted over to different chromosomes or for which the conversion failed. The number of virtually genotyped and imputed variants for each combination of cohort, genotyping array, and imputation panel is summarized in **Table S3**.

#### True phenotype

We simulated 300 true phenotypes that resemble observed complex trait genetic architecture and phenotypic heterogeneity across cohorts. Based on previous literature, we set parameters as follows: 1) 50% of 1 Mb loci contain a true causal variant^94^; 2) probability of being causal is proportional to functional enrichments of variant consequences (pLoF, missense, synonymous, 5’/3’ UTR, promoter, cis-regulatory region, and non-genic) for fine-mapped variants as estimated in a previous complex trait fine-mapping study^17^; 3) per-allele causal effect sizes have a variance proportional to [2*p*(1 − *p*)], where *p* represents a maximum MAF across the three ancestries (AFR, EAS, and EUR) and α is set to be –0.38 (ref. ^52^); and 4) total SNP-heritability *h*^2^ for chromosome 3 equals 0.03 (ref. ^53^). For the sake of simplicity, we randomly draw a single true causal variant per locus because ABF assumes a single causal variant^31,32^. We draw true causal variants from 1,150,893 non-ambiguous single-nucleotide variants in 1000GP3 that showed MAF > 0.01 in at least one of the three ancestries (AFR, EAS, or EUR) and were not located within conversion-unstable positions (CUP)^54^ between the human genome builds GRCh37 and GRCh38. To mimic phenotypic heterogeneity across cohorts in real-world meta-analysis (due to e.g., different ascertainment, measurement error, or true effect size heterogeneity), we introduced cross-cohort genetic correlation of true effect sizes *r*_*g*_ which is set to be one of 1, 0.9, or 0.5. For a true causal variant *j*, true causal effect sizes *βj* across cohorts were randomly drawn from *β*_*j*_∼*MVN*(0, ∑) where diagonal elements of ∑ were set to be 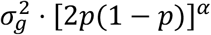 and off-diagonal elements of ∑ were set to be 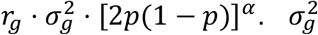 was determined by 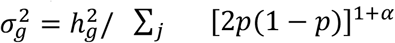. For each cohort, true phenotype *y* was computed via ***y*** = ***Xβ*** + *ϵ* where ***X*** is the above true genotype matrix from HAPGEN2 and 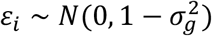 i.i.d. We simulated 100 true phenotypes for each of *r*_*g*_= 1, 0.9, and 0.5, respectively.

#### GWAS

For each combination of phenotype, cohort, genotyping chip, and imputation panel, we conducted GWAS via a standard linear regression as implemented in PLINK 2.0 using imputed dosages. For covariates, we included top 10 principal components that were calculated based on true genotypes after restricting to unrelated samples. We only used LD-pruned variants with MAF > 0.01 for PCA.

#### Meta-analysis

To simulate meta-analyses that resemble real-world settings, we generated multiple *configurations* of the above GWAS results to meta-analyze across 10 independent cohorts. Briefly, we chose configurations based on the following settings: 1) 10 EUR cohorts are genotyped and imputed using the same genotyping array (one of GSA, MEGA, or Omni2.5) and the same imputation panel (one of 1000GP3, HRC, TOPMed, or TOPMed-liftover); 2) 10 cohorts consisting of multiple ancestries (9 EUR + 1 AFR/EAS cohorts or 8 EUR + 1 AFR + 1 EAS cohorts), with all cohorts genotyped and imputed using the same array (Omni2.5) and the same panel (1000GP3); 3) 10 EUR or multi-ancestry cohorts are genotyped using the same array (Omni2.5) but imputed using different panels across cohorts; 4) 10 EUR or multi-ancestry cohorts are imputed using the same panel (1000GP3) but genotyped using different arrays across cohorts; 5) 10 EUR or multi-ancestry cohorts are genotyped and imputed using different arrays and panels across cohorts. For settings 3–5, we randomly draw a combination of a genotyping array and an imputation panel for each cohort five times each for 10 EUR and multi-ancestry cohorts. In total, we generated 45 configurations as summarized in **Table S4**.

For each configuration, we conducted a fixed-effect meta-analysis based on inverse-variance weighted betas and standard errors using a modified version of PLINK 1.9 (https://github.com/mkanai/plink-ng/tree/add_se_meta).

#### Fine-mapping

For each meta-analysis, we defined fine-mapping regions based on a 1 Mb window around each genome-wide significant lead variant and applied ABF^31,32^ using prior effect size variance of 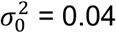. We set a prior variance of effect size to be 0.04 which was taken from Wakefield et al.^31^ and is commonly used in meta-analysis fine-mapping studies^2,7^. We computed posterior inclusion probability (PIP) and 95% credible set (CS) for each locus and evaluated whether true causal variants were correctly fine-mapped.

### The SLALOM method

SLALOM takes GWAS summary statistics and external LD reference as input and predicts whether a locus is suspicious for fine-mapping. SLALOM consists of the following three steps:

#### Locus definition

Consistent with common fine-mapping region definition, we defined loci based on a 1 Mb window around each genome-wide significant lead variant and merged them if they overlapped. We excluded the major histocompatibility complex (MHC) region (chr 6: 25-36 Mb) from analysis due to extensive LD structure in the region.

#### DENTIST-S outlier detection

For each variant in a locus, we computed DENTIST-S statistics using equation (1) based on the assumption of a single causal variant. DENTIST-S P-values (*P*_DENTIST-S_) were computed using the *χ*^2^ distribution with 1 degree of freedom. We applied ABF^31,32^ using prior effect size variance of 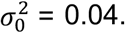 and used the lead PIP variant (the variant with the highest PIP) as an approximation of the causal variant in the locus. To retrieve correlation *r* among the variants, we used publicly available LD matrices from gnomAD^57^ v2 as external LD reference for African, Admixed American, East Asian, Finnish, and non-Finnish European populations. When multiple populations exist, we computed a sample-size-weighted average of *r*^2^ using per-variant sample sizes for each population as previously suggested^81^. We excluded variants without *r*^2^ available in gnomAD from the analysis. Since gnomAD v2 LD matrices are based on the human genome assembly GRCh37, variants were lifted over to GRCh38 if the input summary statistics were based on GRCh38.

We determined DENTIST-S outlier variants using two thresholds: 1) *r*^2^ > *ρ* to the lead and 2) *P*_DENTIST-S_ < *τ*. The thresholds *ρ* and *τ* were set to *ρ* = 0.6 and *τ* = 1.0 × 10^−4^ based on the training in simulations as described below.

#### Suspicious loci prediction

We predicted whether a locus is suspicious or non-suspicious for fine-mapping based on the number of DENTIST-S outlier variants in the locus > *κ*. To determine the best-performing thresholds (*ρ, τ*, and *κ*), we used loci with maximum PIP > 0.9 in the simulations for training. Positive conditions were defined as whether a true causal variant in a locus is 1) a lead PIP variant, 2) in 95% CS, and 3) in 99% CS. We computed AUROC across different thresholds (*ρ* = 0, 0.1, 0.2, …, 0.9; –log_10_ *τ* = 0, 0.5, 1, …, 10; and *κ* = 0, 1, 2, …) and chose *ρ* = 0.6, *τ* = 1.0 × 10^−4^, and *κ* = 0 that showed the highest AUROC for all the aforementioned positive conditions. Using all the loci in the simulations, we then evaluated fine-mapping miscalibration (defined as mean PIP – fraction of true causal variants) at different PIP thresholds in suspicious and non-suspicious loci and decided to only apply SLALOM to loci with maximum PIP > 0.1 owing to relatively lower miscalibration and specificity of SLALOM at lower PIP thresholds.

### GWAS Catalog analysis

We retrieved full GWAS summary statistics publicly available on the GWAS Catalog^48^. Out of 33,052 studies from 5,553 publications registered at the GWAS Catalog (as of January 12, 2022), we selected 467 studies from 96 publications that have 1) full harmonized summary statistics preprocessed by the GWAS Catalog with non-missing variant ID, marginal beta, and standard error columns, 2) a discovery sample size of more than 10,000 individuals, 3) African (including African American, Afro-Caribbean, and Sub-Saharan African), admixed American (Hispanic and Latin American), East Asian, or European samples based on their broad ancestral category metadata, 4) at least one genome-wide significant association (*P* < 5.0 × 10^−8^), and 5) our manual annotation as a meta-analysis rather than a single-cohort study (**Table S6**). We applied SLALOM to the 467 summary statistics and identified 35,864 genome-wide significant loci (based on 1 Mb window around lead variants), of which 28,925 loci with maximum PIP > 0.1 were further classified into suspicious and non-suspicious loci. Since per-variant sample sizes were not available, we used overall sample sizes of each ancestry (African, Admixed American, East Asian, and European) to calculate the weighted-average of *r*^2^. All the variants were harmonized into the human genome assembly GRCh38 by the GWAS Catalog.

### GBMI analysis

We used meta-analysis summary statistics of 14 disease endpoints from the GBMI (**Table S8**). These meta-analyses were conducted using up to 1.8 million individuals across 18 biobanks for discovery, representing six different genetic ancestry groups (approximately 33,000 African, 18,000 Admixed American, 31,000 Central and South Asian, 341,000 East Asian, 1.4 million European, and 1,600 Middle Eastern individuals). Detailed procedures of the GBMI meta-analyses were described in the GBMI flagship publication^10^.

Across the 14 summary statistics, we used 489 out of 500 genome-wide significant loci (*P* < 5.0 × 10^−8^; 1 Mb window around each lead variant, as defined in the GBMI flagship publication^10^), excluding 11 loci that overlap with the MHC region. We applied SLALOM to 422 loci with maximum PIP > 0.1 based on the ABF fine-mapping and predicted whether they were suspicious or non-suspicious for fine-mapping. We used per-variant sample sizes of each ancestry (African, Admixed American, East Asian, Finnish, and non-Finnish European) to calculate the weighted-average of *r*^2^. Since gnomAD LD matrices were not available for Central and South Asian and Middle Eastern, we did not use their sample sizes for the calculation. All the variants were processed on the human genome assembly GRCh38.

### Fine-mapping results of complex traits and *cis*-eQTL

We retrieved our previous fine-mapping results for 1) complex traits in large-scale biobanks (BBJ^58^, FinnGen^20^, and UKBB^19^ Europeans)^16,17^ and 2) *cis*-eQTLs in GTEx^59^ v8 and eQTL Catalogue^60^. Briefly, we conducted multiple-causal-variant fine-mapping (FINEMAP^21,22^ and SuSiE^23^) of complex trait GWAS (# unique traits = 148) and *cis*-eQTL gene expression (# unique tissues/cell-types = 69) using summary statistics and in-sample LD. Detailed fine-mapping methods are described elsewhere^16,17^.

In this study, we collected 1) high-PIP GWAS variants that achieved PIP > 0.9 for any traits in any biobank and 2) high-PIP *cis*-eQTL variants that acheived PIP > 0.9 for any gene expression in any tissues/cell-types. All the variants were originally processed on the human genome assembly GRCh37 and lifted over to the GRCh38 for comparison.

#### Additional fine-mapping results

To compare with the GBMI meta-analyses, we additionally conducted multi-causal-variant fine-mapping of four additional endpoints (gout, heart failure, thyroid cancer, and venous thromboembolism) that were not fine-mapped in our previous study^16,17^. We used exactly the same fine-mapping pipeline (FINEMAP^21,22^ and SuSiE^23^) as described previously^16,17^. For UKBB Europeans, to use the exact same samples that contributed to the GBMI, we used individuals of European ancestry (*n* = 420,531) as defined in the Pan-UKBB project (https://pan.ukbb.broadinstitute.org), instead of those of “white British ancestry” (*n* = 361,194) used in our previous study^16,17^.

### Enrichment analysis of likely causal variants

To validate SLALOM performance, we asked whether suspicious and non-suspicious loci were enriched for having likely causal variants as a lead PIP variant, and for containing them in the 95% and 99% CS. We defined likely causal variants using 1) nonsynonymous coding variants, *i*.*e*., pLoF and missense variants annotated^95^ by the Ensembl Variant Effect Predictor (VEP) v101 (using GRCh38 and GENCODE v35), 2) the high-PIP (> 0.9) complex trait fine-mapped variants, and 3) the high-PIP (> 0.9) *cis*-eQTL fine-mapped variants from our previous studies as described above.

We estimated enrichment for suspicious and non-suspicious loci as a relative risk (*i*.*e*., a ratio of proportion of variants) between being in suspicious/non-suspicious loci and having the annotated likely causal variants as a lead PIP variant (or containing them in the 95% or 99% CS). That is, a relative risk = (proportion of non-suspicious loci having the annotated variants as a lead PIP variant) / (proportion of suspicious loci having the annotated variants as a lead PIP variant). We computed 95% confidence intervals using bootstrapping.

### Comparison of fine-mapping results between the GBMI and individual biobanks

To directly compare with fine-mapping results from the GBMI meta-analyses, we used our fine-mapping results of nine disease endpoints (asthma^64^, COPD^64^, gout, heart failure^73^, IPF^62^, primary open angle glaucoma^74^, thyroid cancer, stroke^75^, and venous thromboembolism^76^) in BBJ^58^, FinnGen^20^, and UKBB^19^ Europeans that were also part of the GBMI meta-analyses for the same traits. For comparison, we computed the maximum PIP for each variant and the minimum size of 95% CS across BBJ, FinnGen, and UKBB. We restricted the 95% CS in biobanks to those that contain the lead variants from the GBMI. We defined the PIP difference between the GBMI and individual biobanks as ΔPIP = PIP (GBMI) – the maximum PIP across the biobanks.

We conducted functional enrichment analysis to compare between the GBMI meta-analysis and individual biobanks because unbiased comparison of PIP requires conditioning on likely causal variants independent of the fine-mapping results, and functional annotations have been shown to be enriched for causal variants. Using functional categories (coding [pLoF, missense, and synonymous], 5’/3’ UTR, promoter, and CRE) from our previous study^16,17^, we estimated functional enrichments of variants in each functional category based on 1) top PIP rankings and 2) ΔPIP bins. Since fine-mapping PIP in the GBMI meta-analysis can be miscalibrated, we performed a comparison based on top PIP rankings to assess whether the ordering given by GBMI PIPs is more informative than the ordering given by the biobanks. For the top PIP rankings, we took the top 0.5%, 0.1%, and 0.05% variants based on the PIP rankings in the GBMI and individual biobanks. We computed enrichment as a relative risk = (proportion of top < Δ PIP variants in the GBMI that are in the annotation) / (proportion of top X% PIP variants in the individual biobanks that are in the annotation). For ΔPIP bins, we defined three bins using different thresholds (*θ* = 0.01, 0.05, and 0.1): 1) decreased PIP bin, ΔPIP < –*θ*, 2) null bin, –*θ* ≤ Δ PIP ≤*θ*, and 3) increased PIP bin, *θ* < ΔPIP. We computed enrichment as a relative risk = (proportion of variants in the decreased/increased PIP bin that are in the annotation) / (proportion of variants in the null PIP bin). We combined coding, UTR, and promoter categories for this analysis due to the limited number of variants for each bin.

## Global Biobank Meta-analysis Initiative

Wei Zhou^1,2,3^, Masahiro Kanai^1,2,3,4,5^, Kuan-Han H Wu^6^, Humaira Rasheed^7,8,9^, Kristin Tsuo^1,2,3^, Jibril B Hirbo^10,11^, Ying Wang^1,2,3^, Arjun Bhattacharya^12^, Huiling Zhao^9^, Shinichi Namba^5^, Ida Surakka^13^, Brooke N Wolford^6,7^, Valeria Lo Faro^14,15,16^, Esteban A Lopera-Maya^17^, Kristi Läll^18^, Marie-Julie Favé^19^, Juulia J Partanen^20^, Sinéad B Chapman^2,3^, Juha Karjalainen^1,2,3,20^, Mitja Kurki^1,2,3,20^, Mutaamba Maasha ^1,2,3,20^, Ben M Brumpton^7,21,22^, Sameer Chavan^23^, Tzu-Ting Chen^24^, Michelle Daya^23^, Yi Ding^25,12^, Yen-Chen A Feng^26^, Lindsay A Guare^27^, Christopher R Gignoux^23^, Sarah E Graham^13^, Whitney E Hornsby^13^, Nathan Ingold^28,29^, Said I. Ismail^30^, Ruth Johnson^31,12^, Triin Laisk^18^, Kuang Lin^32^, Jun Lv^33^, Iona Y Millwood^32,34^, Sonia Moreno-Grau^35^, Kisung Nam^36^, Priit Palta^18,20^, Anita Pandit^37^, Michael H Preuss^38^, Chadi Saad^30^, Shefali S Setia^39^, Unnur Thorsteinsdottir^40^, Jasmina Uzunovic^19^, Anurag Verma^41,42^, Matthew Zawistowski^37^, Xue Zhong^10,11^, Nahla Afifi^43^, Kawthar M. Al-Dabhani^43^, Asma Al Thani^43^, Yuki Bradford^27^, Archie Campbell^44^, Kristy Crooks^23^, Geertruida H de Bock^45^, Scott M Damrauer^46,27,42^, Nicholas J Douville^47,48^, Sarah Finer^49^, Lars G Fritsche^37^, Eleni Fthenou^43^, Gilberto Gonzalez-Arroyo^35,50^, Christopher J Griffiths^49^, Yu Guo^51^, Karen A Hunt^52^, Alexander Ioannidis^35,53^, Nomdo M Jansonius^14^, Takahiro Konuma^5,54^, Ming Ta Michael Lee^35^, Arturo Lopez-Pineda^35,50^, Yuta Matsuda^55^, Riccardo E Marioni^44^, Babak Moatamed^35^, Marco A Nava-Aguilar^35,50^, Kensuke Numakura^55^, Snehal Patil^37^, Nicholas Rafaels^23^, Anne Richmond^56^, Agustin Rojas-Muñoz^35^, Jonathan A Shortt^23^, Peter Straub^10,11^, Ran Tao^57,11^, Brett Vanderwerff^37^, Manvi Vernekar^55^, Yogasudha Veturi^27^, Kathleen C Barnes^23^, Marike Boezen^45^‡, Zhengming Chen^32,34^, Chia-Yen Chen^58^, Judy Cho^38^, George Davey Smith^9,59^, Hilary K Finucane^1,3,2^, Lude Franke^17^, Eric R Gamazon^10,11,60^, Andrea Ganna^1,2,20^, Tom R Gaunt^9,59^, Tian Ge^61,62^, Hailiang Huang^1,2^, Jennifer Huffman^63^, Nicholas Katsanis^35^, Jukka T Koskela^20^, Clara Lajonchere^64,65^, Matthew H Law^28,29^, Liming Li^33^, Cecilia M Lindgren^66^, Ruth JF Loos^38,67^, Stuart MacGregor^28^, Koichi Matsuda^68^, Catherine M Olsen^28^, David J Porteous^44^, Jordan A Shavit^69^, Harold Snieder^45^, Tomohiro Takano^55^, Richard C Trembath^70^, Judith M Vonk^45^, David C. Whiteman^28^, Stephen J Wicks^23^, Cisca Wijmenga^17^, John Wright^71^, Jie Zheng^9^, Xiang Zhou^37^, Philip Awadalla^19,72^, Michael Boehnke^37^, Carlos D Bustamante^35,53,73^, Nancy J Cox^10,11^, Segun Fatumo^74,75,76^, Daniel H Geschwind^77,64,78^, Caroline Hayward^56^, Kristian Hveem^7,21^, Eimear E Kenny^79^, Seunggeun Lee^36^, Yen-Feng Lin^24,80,81^, Hamdi Mbarek^30^, Reedik Mägi^18^, Hilary C Martin^82^, Sarah E Medland^28^, Yukinori Okada^5,83,84,85,86^, Aarno V Palotie^1,2,20^, Bogdan Pasaniuc^12,77,87,25,64^, Daniel J Rader^27,41^, Marylyn D Ritchie^27^, Serena Sanna^88,17^, Jordan W Smoller^61^, Kari Stefansson^40^, David A van Heel^52^, Robin G Walters^32,34^, Sebastian Zöllner^37^, Biobank of the Americas, Biobank Japan Project, BioMe, BioVU, CanPath - Ontario Health Study, China Kadoorie Biobank Collaborative Group, Colorado Center for Personalized Medicine, deCODE Genetics, Estonian Biobank, FinnGen, Generation Scotland, Genes & Health Research Team, LifeLines, Mass General Brigham Biobank, Michigan Genomics Initiative, National Biobank of Korea, Penn Medicine BioBank, Qatar Biobank, The QSkin Sun and Health Study, Taiwan Biobank, The HUNT Study, UCLA ATLAS Community Health Initiative, Uganda Genome Resource, UK Biobank, Alicia R Martin^1,2,3^, Cristen J Willer^13,6,89^*, Mark J Daly^1,2,3,20^*, Benjamin M Neale^1,2,3^*

‡Deceased

*These authors jointly supervised the initiative

^1^Analytic and Translational Genetics Unit, Department of Medicine, Massachusetts General Hospital, Boston, MA, USA, ^2^Stanley Center for Psychiatric Research, Broad Institute of MIT and Harvard, Cambridge, MA, USA, ^3^Program in Medical and Population Genetics, Broad Institute of MIT and Harvard, Cambridge, MA, USA, ^4^Department of Biomedical Informatics, Harvard Medical School, Boston, MA, USA, ^5^Department of Statistical Genetics, Osaka University Graduate School of Medicine, Suita 565-0871, Japan, ^6^Department of Computational Medicine and Bioinformatics, University of Michigan, Ann Arbor, MI, USA, ^7^K.G. Jebsen Center for Genetic Epidemiology, Department of Public Health and Nursing, NTNU, Norwegian University of Science and Technology, Trondheim, Norway, ^8^Division of Medicine and Laboratory Sciences, University of Oslo, Norway, ^9^MRC Integrative Epidemiology Unit (IEU), Bristol Medical School, University of Bristol, Bristol, UK, ^10^Department of Medicine, Division of Genetic Medicine, Vanderbilt University Medical Center, Nashville, TN, USA, ^11^Vanderbilt Genetics Institute, Vanderbilt University Medical Center, Nashville, TN, USA, ^12^Department of Pathology and Laboratory Medicine, David Geffen School of Medicine, University of California, Los Angeles, Los Angeles, CA, USA, ^13^Department of Internal Medicine, Division of Cardiology, University of Michigan, Ann Arbor, MI, USA, ^14^University of Groningen, UMCG, Department of Ophthalmology, Groningen, the Netherlands, ^15^Department of Clinical Genetics, Amsterdam University Medical Center (AMC), Amsterdam, the Netherlands, ^16^Department of Immunology, Genetics and Pathology, Science for Life Laboratory, Uppsala University, Uppsala, Sweden, ^17^University of Groningen, UMCG, Department of Genetics, Groningen, the Netherlands, ^18^Estonian Genome Centre, Institute of Genomics, University of Tartu, Tartu, Estonia, ^19^Ontario Institute for Cancer Research, Toronto, ON, Canada, ^20^Institute for Molecular Medicine Finland, University of Helsinki, Helsinki, Finland, ^21^HUNT Research Centre, Department of Public Health and Nursing, NTNU, Norwegian University of Science and Technology, Levanger, Norway, ^22^Clinic of Medicine, St. Olavs Hospital, Trondheim University Hospital, Trondheim, Norway, ^23^University of Colorado - Anschutz Medical Campus, Aurora, CO, USA, ^24^Center for Neuropsychiatric Research, National Health Research Institutes, Miaoli, Taiwan, ^25^Bioinformatics Interdepartmental Program, University of California, Los Angeles, Los Angeles, CA, USA, ^26^Division of Biostatistics, Institute of Epidemiology and Preventive Medicine, College of Public Health, National Taiwan University, Taiwan, ^27^Department of Genetics, Perelman School of Medicine, University of Pennsylvania, Philadelphia, PA, USA, ^28^QIMR Berghofer Medical Research Institute, Brisbane, Australia, ^29^Faculty of Health, School of Biomedical Sciences, Queensland University of Technology, Australia, ^30^Qatar Genome Program, Qatar Foundation Research, Development and Innovation, Qatar Foundation, Doha, Qatar, ^31^Department of Computer Science, University of California, Los Angeles, Los Angeles, CA, USA, ^32^Nuffield Department of Population Health, University of Oxford, Oxford, UK, ^33^Department of Epidemiology and Biostatistics, School of Public Health, Peking University Health Science Center, Beijing, China, ^34^MRC Population Health Research Unit, University of Oxford, Oxford, UK, ^35^Galatea Bio Inc., Hialeah, FL, USA, ^36^Graduate School of Data Science, Seoul National University, ^37^Department of Biostatistics and Center for Statistical Genetics, University of Michigan, Ann Arbor, MI, USA, ^38^The Charles Bronfman Institute for Personalized Medicine, Icahn School of Medicine at Mount Sinai, New York, NY, USA, ^39^Department of Pathology and Laboratory Medicine, Perelman School of Medicine, University of Pennsylvania, Philadelphia, PA, USA, ^40^deCODE Genetics/Amgen inc., 101, Reykjavik, Iceland, ^41^Department of Medicine, Perelman School of Medicine, University of Pennsylvania, Philadelphia, PA, USA, ^42^Corporal Michael Crescenz VA Medical Center, Philadelphia, PA, USA, ^43^Qatar Biobank for Medical Research, Qatar Foundation for Education, Science, and Community, Doha, Qatar, ^44^Centre for Genomic and Experimental Medicine, Institute of Genetics and Cancer, University of Edinburgh, Edinburgh, UK, ^45^Department of Epidemiology, University Medical Center Groningen, Groningen, the Netherlands, ^46^Department of Surgery, Perelman School of Medicine, University of Pennsylvania, Philadelphia, PA, USA, ^47^Department of Anesthesiology, Michigan Medicine, Ann Arbor, MI, USA, ^48^Institute of Healthcare Policy & Innovation, University of Michigan, Ann Arbor, MI, USA, ^49^Wolfson Institute of Population Health, Queen Mary University of London, London, UK, ^50^Amphora Health, Morelia, Michoacan, Mexico, ^51^Chinese Academy of Medical Sciences, Beijing, China, ^52^Blizard Institute, Queen Mary University of London, London, UK, ^53^Stanford University School of Medicine, Stanford, CA, USA, ^54^Central Pharmaceutical Research Institute, JAPAN TOBACCO INC., Takatsuki 569-1125, Japan, ^55^Genomelink, Inc., Berkeley, CA, USA, ^56^Medical Research Council Human Genetics Unit, Institute of Genetics and Cancer, University of Edinburgh, Edinburgh, UK, ^57^Department of Biostatistics, Vanderbilt University Medical Center, Nashville, TN, USA, ^58^Biogen, Cambridge, MA, USA, ^59^NIHR Bristol Biomedical Research Centre, Bristol, UK, ^60^MRC Epidemiology Unit, University of Cambridge, Cambridge, UK, ^61^Psychiatric and Neurodevelopmental Genetics Unit, Center for Genomic Medicine, Massachusetts General Hospital, Boston, MA, USA, ^62^Center for Precision Psychiatry, Massachusetts General Hospital, Boston, MA, USA, ^63^Centre for Population Genomics, VA Boston Healthcare System, Boston, MA, USA, ^64^Institute of Precision Health, University of California, Los Angeles, Los Angeles, CA, USA, ^65^Program in Neurogenetics, Department of Neurology, David Geffen School of Medicine, University of California, Los Angeles, Los Angeles, CA, USA, ^66^Big Data Institute, Li Ka Shing Centre for Health Information and Discovery, University of Oxford, Oxford, UK, ^67^Novo Nordisk Foundation Center for Basic Metabolic Research, Faculty of Medicine and Health Sciences, University of Copenhagen, Copenhagen, Denmark, ^68^Department of Computational Biology and Medical Sciences, Graduate school of Frontier Sciences, The University of Tokyo, Tokyo, Japan, ^69^University of Michigan, Department of Pediatrics, Ann Arbor MI 48109, ^70^School of Basic and Medical Biosciences, Faculty of Life Sciences and Medicine, King’s College London, London, UK, ^71^Bradford Institute for Health Research, Bradford Teaching Hospitals National Health Service (NHS) Foundation Trust, Bradford, UK, ^72^Department of Molecular Genetics, University of Toronto, Toronto, ON, Canada, ^73^Chan Zuckerberg Biohub, San Francisco, CA, USA, ^74^The African Computational Genomics (TACG) Research Group, MRC/UVRI and LSHTM, Entebbe, Uganda, ^75^London School of Hygiene & Tropical Medicine London, UK, ^76^Medical Research Council/ Uganda Virus Research Institute/London School of Hygiene and Tropical Medicine (MRC/UVRI/LSHTM) Uganda research unit, Entebbe, Uganda, ^77^Department of Human Genetics, David Geffen School of Medicine, University of California, Los Angeles, Los Angeles, CA, USA, ^78^Department of Neurology, David Geffen School of Medicine, University of California, Los Angeles, Los Angeles, CA, USA, ^79^Institute for Genomic Health, Icahn School of Medicine at Mount Sinai, New York, NY, USA, ^80^Department of Public Health & Medical Humanities, School of Medicine, National Yang Ming Chiao Tung University, Taipei, Taiwan, ^81^Institute of Behavioral Medicine, College of Medicine, National Cheng Kung University, Tainan, Taiwan, ^82^Medical and Population Genomics, Wellcome Sanger Institute, Hinxton, UK, ^83^Department of Genome Informatics, Graduate School of Medicine, the University of Tokyo, Tokyo 113-0033, Japan., ^84^Laboratory of Statistical Immunology, Immunology Frontier Research Center (WPI-IFReC), Osaka University, Suita 565-0871, Japan, ^85^Laboratory for Systems Genetics, RIKEN Center for Integrative Medical Sciences, Yokohama, Japan, ^86^Integrated Frontier Research for Medical Science Division, Institute for Open and Transdisciplinary Research Initiatives, Osaka University, Suita 565-0871, Japan, ^87^Department of Computational Medicine, David Geffen School of Medicine, University of California, Los Angeles, Los Angeles, CA, USA, ^88^Institute for Genetics and Biomedical Research (IRGB), National Research Council (CNR), Cagliari, Italy, ^89^Department of Human Genetics, University of Michigan, Ann Arbor, MI, USA

